# Short-Term Effect of Ambient Meteorological Factors on Hand-Foot-Mouth Disease: An Individual-Level Case-Crossover Study in Jiangsu, China

**DOI:** 10.1101/2025.03.11.25323805

**Authors:** Yifan Tang, Hong Ji, Hongfei Zhu, Xu Wang, Long Bai, Wendong Liu, Kai Wang, Conghua Wen, Ying Wang, Chengxiu Ling, Liguo Zhu

**Author notes:** These authors contributed equally to the work (Yifan Tang, Hong Ji, Hongfei Zhu).

## Abstract

**Background:** Hand, foot, and mouth disease (HFMD) represent a significant public health concern in the Asia-Pacific region, imposing a substantial burden that warrants urgent attention. However, the associations between individual-level exposure to ambient meteorological factors and the HFMD risk remain poorly understood.

**Methods:** Using a time-stratified case-crossover design, we examine the individual-level association between six meteorological factors (temperature, humidity, wind speed, radiation, surface pressure, and precipitation) and HFMD risk. Conditional logistic regression is employed to investigate the relationship between short-term exposure to meteorological factors and HFMD risk, considering lagged effects and adjusting for public holidays and time-varying grid-level HFMD susceptibility. Exposure-response curves are developed using natural cubic splines to model the non-linear associations, and then extreme meteorological exposure under different thresholds is considered.

**Results:** A total of 1,247,970 eligible cases are identified in the study. Each 1-unit increase in exposure to temperature, humidity, wind speed, and precipitation over a 10-day moving average (lag010) is associated with an elevated risk of HFMD, with odds ratios (ORs) of 1.0124 (95% CI: 1.0013, 1.0134), 1.0063 (95% CI: 1.0059, 1.0068), 1.0069 (95% CI: 1.0022, 1.0115), and 1.0081 (95% CI: 1.0069, 1.0092), respectively. In contrast, radiation and surface pressure exhibited a negative association with HFMD, with ORs of 0.9860 (95% CI: 0.9848, 0.9871) and 0.8780 (95% CI: 0.8660, 0.8900) at lag010. As the exposure thresholds for temperature, humidity, wind speed, and precipitation increase, the negative association between the excess magnitude and HFMD risk is strengthened, whereas the associations for radiation and surface pressure are reversed.

**Conclusion:** Our findings indicate that short-term exposure to most meteorological factors, except radiation and surface pressure, is associated with an elevated risk of HFMD, providing valuable insights for developing targeted preventive strategies and public health policies.

## 1. Introduction

Hand, foot, and mouth disease (HFMD) is a common viral infection, particularly affecting children under 5 years of age, with main clinical signs including fever, mouth ulcers, and blisters.^1,2^ While it is generally self-limiting in most cases,^3^ severe instances can lead to fatal neurological and systemic complications, and it remains infectious with the highest mortality rate among Category C infectious diseases implemented in mainland China.^4^ China reports approximately 2 million Hand, Foot, and Mouth Disease (HFMD) cases annually, with the incidence rate varying significantly across regions, ranging from 37.01 to 205.06 per 100,000 individuals.^5^ Given the threat that HFMD poses to children and its persistently high case numbers, it represents a major public health problem in the Asia-Pacific region that cannot be ignored.^6^

Numerous studies have underscored the significant role of meteorological factors in transmitting infectious diseases, such as variations in temperature and precipitation driving the spread of malaria, dengue fever, Lyme disease, and cholera.^7^ These resulting climate-sensitive health burdens necessitated strengthening prevention and control strategies, particularly for infectious diseases, as emphasized in the 2022 Lancet Countdown Report on Population Health and Climate Change.^8^ In the context of HFMD, earlier studies typically considered linear associations between meteorological factors and disease risk. However, these yielded inconsistent findings,^9,10^ suggesting that a more nuanced approach is needed. Recent research has relaxed the assumption of linearity, exploring potential non-linear relationships. For example, studies in Japan and China have demonstrated that HFMD risk increases with moderate temperatures but decreases at extreme temperature thresholds, forming an inverted V-shape curve.^11,12^ Methods such as generalized additive models (GAM), splines, and distributed lag nonlinear models (DLNM) have been used to capture these non-linear patterns, offering greater flexibility in understanding the climate-disease relationship.^13,14^ To account for regional variability in both disease and environmental conditions, the Bayesian spatiotemporal model, which incorporates latent spatial and temporal effects, has been increasingly applied.^15,16^

Note that most aforementioned studies rely on aggregated data at the municipal level, and the potential risk of ecological fallacies is evident.^9,12,15^ Due to the presence of unmeasured individual-level confounders, the associations between meteorological factors and HFMD found at the municipal level may not accurately reflect the true exposure-response relationships at the individual level.^17,18^ To address these limitations, in this study, we employ a self-matched case-crossover design as an alternative modeling approach to examine individual-level associations between meteorological factor exposure and HFMD in the high-resolution context of Jiangsu, China. This design improves upon traditional aggregate-level analyses by accounting for within-individual variability and minimizing potential confounding factors that may be obscured in aggregated data, thereby providing more precise estimates of the exposure-outcome relationship.^17, 19^

This study leverages a large-scale surveillance dataset from the National Infectious Diseases Reporting System (NIDRS) to assess the associations between short-term individual-level exposure to six meteorological factors (temperature, humidity, wind speed, radiation, surface pressure, precipitation) and HFMD using home address-linked gridded meteorological data spanning 15 years, from 2009 to 2023. Additionally, we explore the potential impact of extreme exposures on HFMD risk. To examine risk stratification across subgroups, we further assess effect modifications by sex, age, and diagnostic category.^17^ Finally, the robustness of our findings was demonstrated through several sensitivity analyses, incorporating additional meteorological factors into the DLNM models, evaluating the effects of the COVID-19 pandemic by comparing periods before and after the 2019 outbreak and adjusting for daytime exposure. To the best of our knowledge, this is the first high-resolution, individual-level HFMD epidemiological study with a case-crossover design examining the associations between exposure to key meteorological factors and HFMD risk.

## 2. Methods

### 2.1 HFMD surveillance data

Data on daily individual cases of HFMD over the period between 1 January 2009 and 31 December 2023 in Jiangsu were obtained from the National Infectious Diseases Reporting System (NIDRS). For each case, information regarding the date of birth, sex, onset date, residential address, and diagnostic type are extracted from the surveillance system. Cases are excluded from the analysis under the following conditions: (1) off-site medical treatment where the patient’s registered domicile was outside the province (N = 20,036, 1.2%), and (2) cases with incomplete or imprecise residential address data lower than the precision of 9 km × 9 km of the meteorological exposure factors (N = 310,696, 19.68%). This results in a final analytical sample of 1,247,970 eligible HFMD cases.

### 2.2 Meteorology exposure assessment

Using meteorological grid data from the European Centre for Medium-Range Weather Forecasts (ECMWF) ERA5 Land Reanalysis (spatial resolution: 9 km × 9 km), we obtained the daily 24-hour average values of temperature, humidity, wind speed, radiation, surface pressure, and precipitation (see detailed descriptions in Table A.1) in Jiangsu province during the study period, and assessed residential exposure to these meteorological conditions during the case days and corresponding control days for each HFMD case.

### 2.2 Study design

We estimate the associations between exposure to meteorological factors and HFMD using a self-matched, time-stratified case-crossover design, a standard epidemiological approach for evaluating the transient effects of environmental exposures on various health outcomes.^20,21^ In this design, each participant serves as their own control, which helps mitigate potential bias arising from long-term trends, seasonality, and unmeasured time-varying confounders that are unlikely to change over short periods, as well as from time-invariant individual-level confounders such as genotype, sex, age, life/dietary status, socioeconomic status, and vaccination.^19^

Under this design, stratification occurs based on patient, day of the week within year-month in the same location. Controls are chosen bidirectionally, both before and after the case, at a ratio of 1:3 or 1:4. The number of controls for a specific case depends on the number of days of the week within a given month.^17^ Each participant’s exposure on the hazard day (the case day) is then compared with exposures on the matched reference days (control days). For example, if a patient falls ill on June 12, 2023 (Monday), June 2023 is defined as the time stratum, June 12, 2023, as the case day, and all other Mondays in June (i.e., June 5, 19, and 26) as corresponding control days. Given that each case day can be matched with 3 or 4 control days, our analysis includes 1,247,970 case days and 4,259,379 control days.

### 2.4 Statistical method

Conditional logistic regression models are used to assess the association between HFMD risk and short-term exposures to each of the six meteorological factors (temperature, humidity, wind speed, radiation, surface pressure, and precipitation). The following formula depicts the individual-level time-stratified case-crossover design^22^:

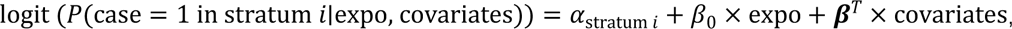

where each stratum consists of one case (coded as 1) and three or four matched controls (coded as 0 for the corresponding controls), *P*(case = 1 in stratum *i*|expo, covariates) is the conditional probability of being a case in the *i* -th stratum after being adjusted for meteorological factor exposure and covariates, *i*=1, 2,…, N = 1,247,970, the total number of HFMD cases. The term *α*_stratum *i*_ refers to the intercept of stratum *i*, and expo represents the meteorological factor exposure with coefficient *β'*_0_, covariates are used to adjust for in the models, including holiday effect and time-varying grid-level HFMD susceptibility with coefficient vector ***β***.

To further specifically explore the impact of extreme meteorological exposure on HFMD risk, we use the following modified model:

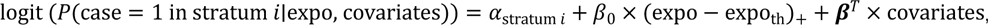

where expo_th_ denotes the exposure threshold, which is set as the 85th, 90th, and 95th percentiles of the exposure distribution.

The holiday effect is accounted for by introducing a binary holiday variable (1: New Year’s Day, the Spring Festival, the Qing Ming Festival, the Labor Day, the Dragon-Boat Festival, the Mid-Autumn Festival, the National Day, Winter vacation, Summer vacation, and weekends; 0: non-holiday). Spatiotemporal clustering of HFMD cases, measured as the total number of HFMD cases from the day of symptom onset up to the previous 7 lag days within a grid for each case, is employed as a proxy for time-varying grid-level HFMD susceptibility. By considering the epidemic phase within each grid, this approach helps distinguish the direct effects of meteorological factors from the influence of ongoing transmission dynamics, thereby reducing potential estimation bias.

Given HFMD’s incubation period of 2-10 days,^1^ both single-lag models (from lag 0 to lag 10) and a moving average lag model (lag 01 to lag 010) are applied to explore the characteristics of lag effects. We also develop nonlinear exposure-response curves to estimate nonlinear associations between specific atmospheric factor exposures and HFMD. Natural cubic splines with two interior knots located at the 25th and 75th percentiles model the exposure-response curve for each meteorological factor.^19,23^

### 2.4 Stratified and sensitivity analyses

We perform subgroup analyses by sex,^24^ age^25^, and diagnosis type.^17^ To evaluate the statistical significance of differences in the effect of meteorological exposure on HFMD across subgroups, *P*-values are computed for the interaction terms between each meteorological factor and subgroup characteristic (e.g., temperature by sex).

Several sensitivity tests are conducted to examine the robustness of our primary findings. First, we incorporate each additional meteorological factor separately into the model to fit a two-meteorological-factor model. Second, a DLNM is used to assess the association between meteorological exposure and HFMD onset, estimating both nonlinear exposure-response and delayed effects with bidimensional cross-basis functions. Third, we conduct analyses separately for cases with disease onset during 2009-2019 and 2020-2023 to assess the potential impact of the pandemic on the findings. Finally, acknowledging that children are not typically exposed to the external environment during nighttime, the average exposure to meteorological factors between 7 a.m. and 8 p.m. each day is used in the sensitivity analysis.

All statistical analyses were conducted using R software (version 4.4.1). The study is approved by the Xi’an Jiaotong-Liverpool University Research Ethics Committee (00100000 89620241213145739).

## 3. Methods

### 3.1 Study sample size

Figure 1 illustrates the distribution of HFMD cases across Jiangsu Province, China, from 2009 to 2023. The geographical distribution is predominantly concentrated in southern Jiangsu. Trend analysis (see Figure S1) over the study period did not reveal any significant temporal trends in the number of HFMD cases. However, seasonal analysis indicates a distinct peak in case numbers during the summer and autumn months, as evidenced by recurrent surges in cases during these periods.

**Figure 1:**
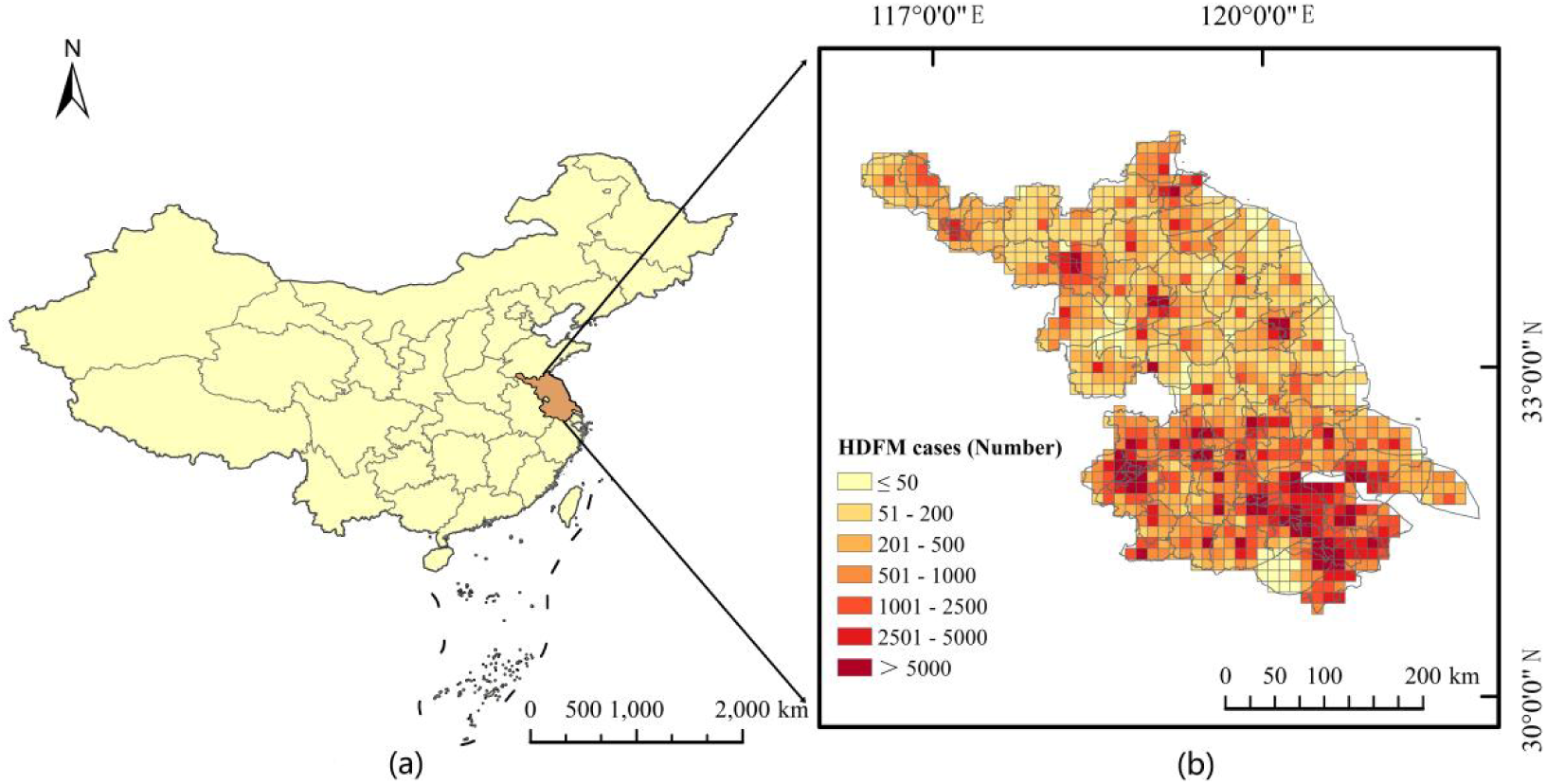
(a) The geographical location and administrative division boundary map of Jiangsu province, China, obtained from the primary geographic database in the National Catalogue Service For Geographic Information of China. (b) Spatial distribution of HFMD cases in Jiangsu Province from 2009 to 2023.

Table 1 displays the characteristics of HFMD cases. There are 1,247,970 case days and 4,259,379 control days for the analysis, and the mean age of cases is 3 years old. A majority of cases are children under 3 years old (657,060, 52.65%) and males (748,689, 59.99%). In addition, there are 740,722 (59.35%) laboratory confirmed and 437,371 (35.05%) clinically diagnosed, while 69,393 (5.56%) confirmed.

**Table 1:** Characteristics of HFMD cases in Jiangsu, China, 2009-2023 for 1,247,970 participants.

| Characteristic |  | N (%) |
| --- | --- | --- |
| Case days (N) |  | 1,247,970 |
| Control days (N) |  | 4,259,379 |
| Age (y) |  |  |
|  | [0, 1) | 113,679 (9.11%) |
|  | [1, 2) | 317,638 (25.42%) |
|  | [2, 3) | 225,743 (18.09%) |
|  | [3, 4) | 231,180 (18.52%) |
|  | [4, 5) | 167,815 (13.45%) |
|  | [5, 6) | 85,331 (6.84%) |
|  | [6, +∞) | 106,584 (8.54%) |
| Sex |  |  |
|  | Male | 748,689 (59.93%) |
|  | Female | 499,281 (40.07%) |
| Diagnostic type |  |  |
|  | Laboratory confirmed | 740,722 (59.35%) |
|  | Clinically confirmed | 437,371 (35.05%) |
|  | Confirmed | 69,393 (5.56%) |

### 3.2 Vulnerability covariate distribution

From 2009 to 2023, the mean exposure to ambient temperature, humidity, wind speed, radiation, surface pressure, and precipitation in each grid is 15.88 ℃, 72.91%, 71.55 m/s, 10.01 mj/m^2^, 101.49 kPa, and 1.57 mm, respectively (see Table 2). The spatial distribution of the 90\% thresholds for these meteorological exposures across each grid in Jiangsu Province, China, from 2009 to 2023 is shown in Figure 2. Figures S1 and S2 illustrate the 85% and 95% thresholds, respectively. Significant spatial variation in these thresholds is observed across the grids. The thresholds for temperature, humidity, and precipitation tend to decrease from south to north, while those for wind speed and radiation are significantly higher in the coastal area. Additionally, the surface pressure thresholds decrease as one moves inland from the coast. Figure S1 shows the time-series plots. Spearman correlations between the six meteorological exposure factors are summarized in Table S1. Temperature is moderately associated with humidity and radiation but strongly correlates negatively with surface pressure. Humidity shows a moderate positive association with precipitation and weak associations with other variables, while wind speed exhibits negligible associations across the board. Radiation is moderately negatively associated with both surface pressure and precipitation, and surface pressure is negatively associated with all variables, most strongly with temperature.

**Figure 2:**
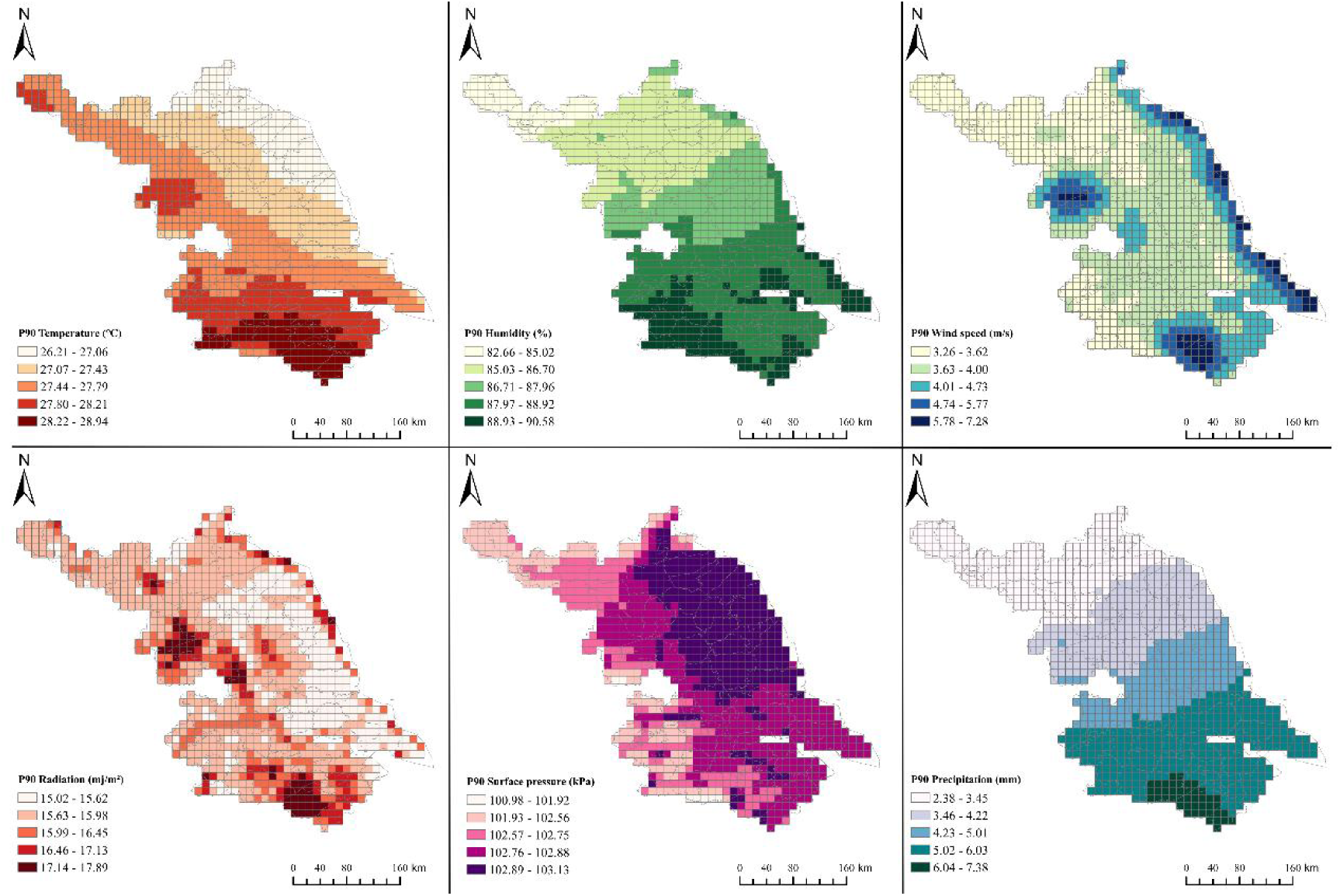
Spatial distribution of meteorological factors 90th percentile thresholds in Jiangsu, China, 2009-2023.

**Table 2:** Descriptive statistics of meteorological factors in Jiangsu, China, 2009-2023.

| Covariate | Mean | (Min, Max) | Sd |
| --- | --- | --- | --- |
| Temperature (°C) | 15.88 | (−13.24, 33.58) | 9.33 |
| Humidity (%) | 72.91 | (14.24,99.35) | 13.14 |
| Wind speed (m/s) | 71.55 | (0.00,16.15) | 1.23 |
| Radiation (mj/m<sup>2</sup>) | 10.01 | (0.28,21.05) | 4.44 |
| Surface pressure (kPa) | 101.49 | (96.75,104.39) | 0.97 |
| Precipitation (mm) | 1.57 | (0.00,187.52) | 4.42 |

### 3.3 Meteorological factors and HFMD risk

The results of the conditional logistic regression models examining the associations between meteorological exposures and HFMD are shown in Figures 3 and 4. In the single-lag models, significant positive associations between meteorological exposures and HFMD are observed for temperature, humidity, wind speed, and precipitation, with effects noted lag 4 to lag 10. Conversely, most single-lag models for radiation and surface pressure indicate a negative association with HFMD risk. Specifically, each 11 mj/m^2^ increase in radiation and each 1 kPa increase in surface pressure at lag 6 are associated with a 0.73% (OR = 0.9927, 95% CI: 0.9922, 0.9932) and 7.79% (OR = 0.9221, 95% CI: 0.9175, 0.9267) reduction in HFMD risk, respectively. Similar trends emerged in the moving average lag models, though with some variation. These models reveal significant short-term associations between meteorological exposures and HFMD, with the strongest associations identified for temperature (OR = 1.0124, 95% CI: 1.0013, 1.0134), humidity (OR = 1.0063, 95% CI: 1.0059, 1.0068), wind speed (OR = 1.0069, 95% CI: 1.0022, 1.0115), radiation (OR = 0.9860, 95% CI: 0.9848, 0.9871), surface pressure (OR = 0.9089, 95% CI: 0.9010, 0.9169), and precipitation (OR = 1.0081, 95% CI: 1.0069, 1.0092), with the most pronounced effects observed at the lag 010 days.

**Figure 3:**
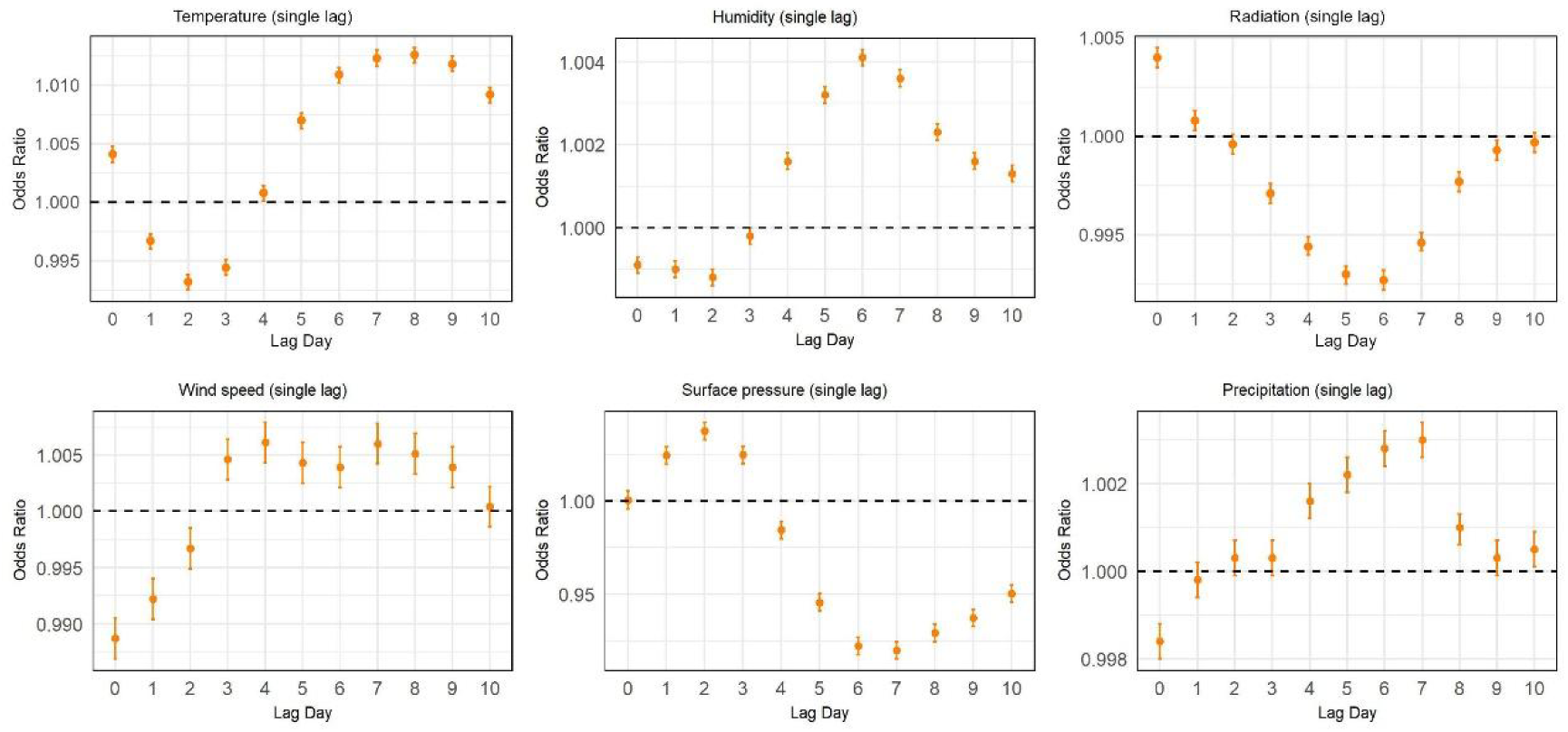
Associations between acute exposures to six meteorological factors and HFMD at different single lag days in Jiangsu, China, 2009-2023.

**Figure 4:**
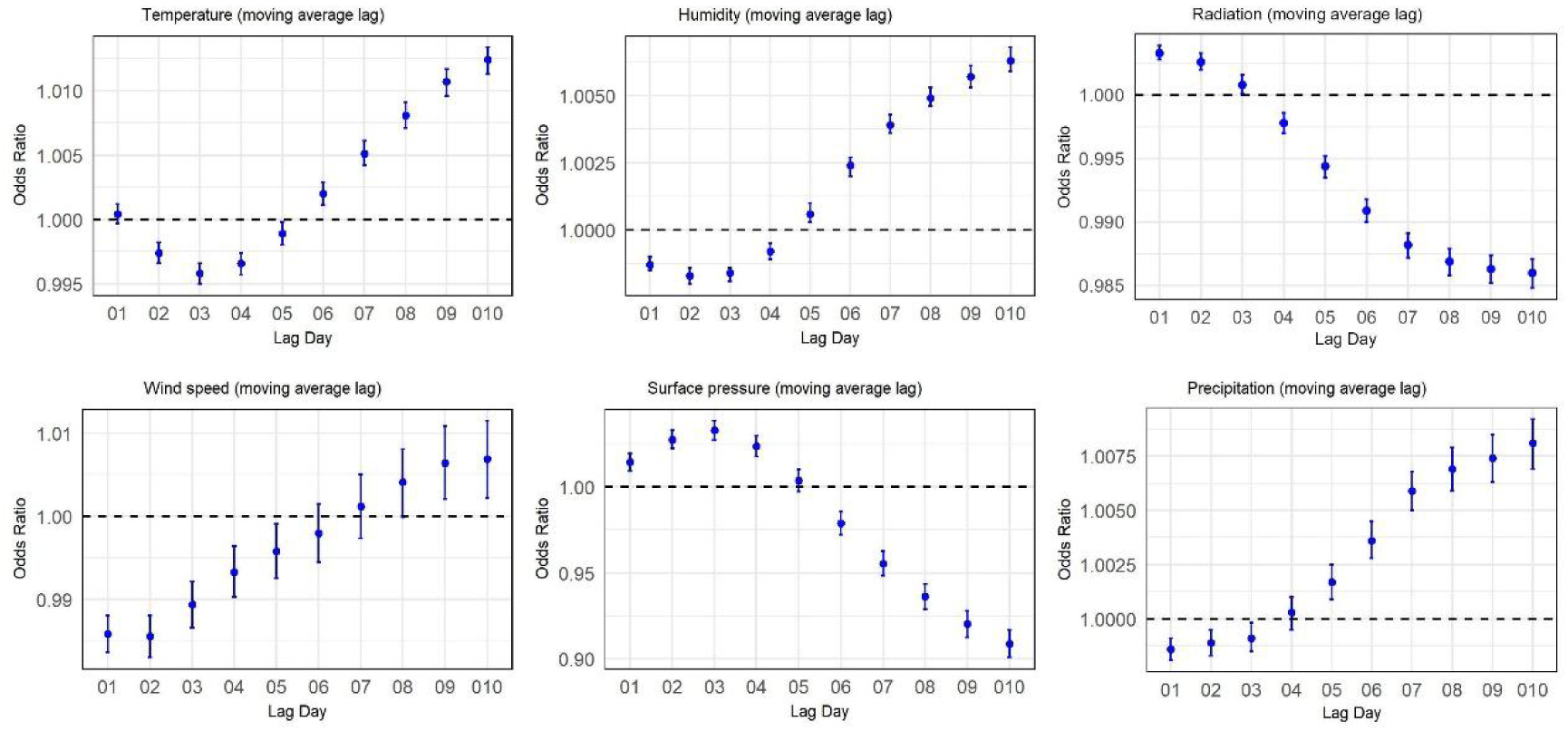
Associations between acute exposures to six meteorological factors and HFMD at different moving average lag days in Jiangsu, China, 2009-2023.

**Figure 5:**
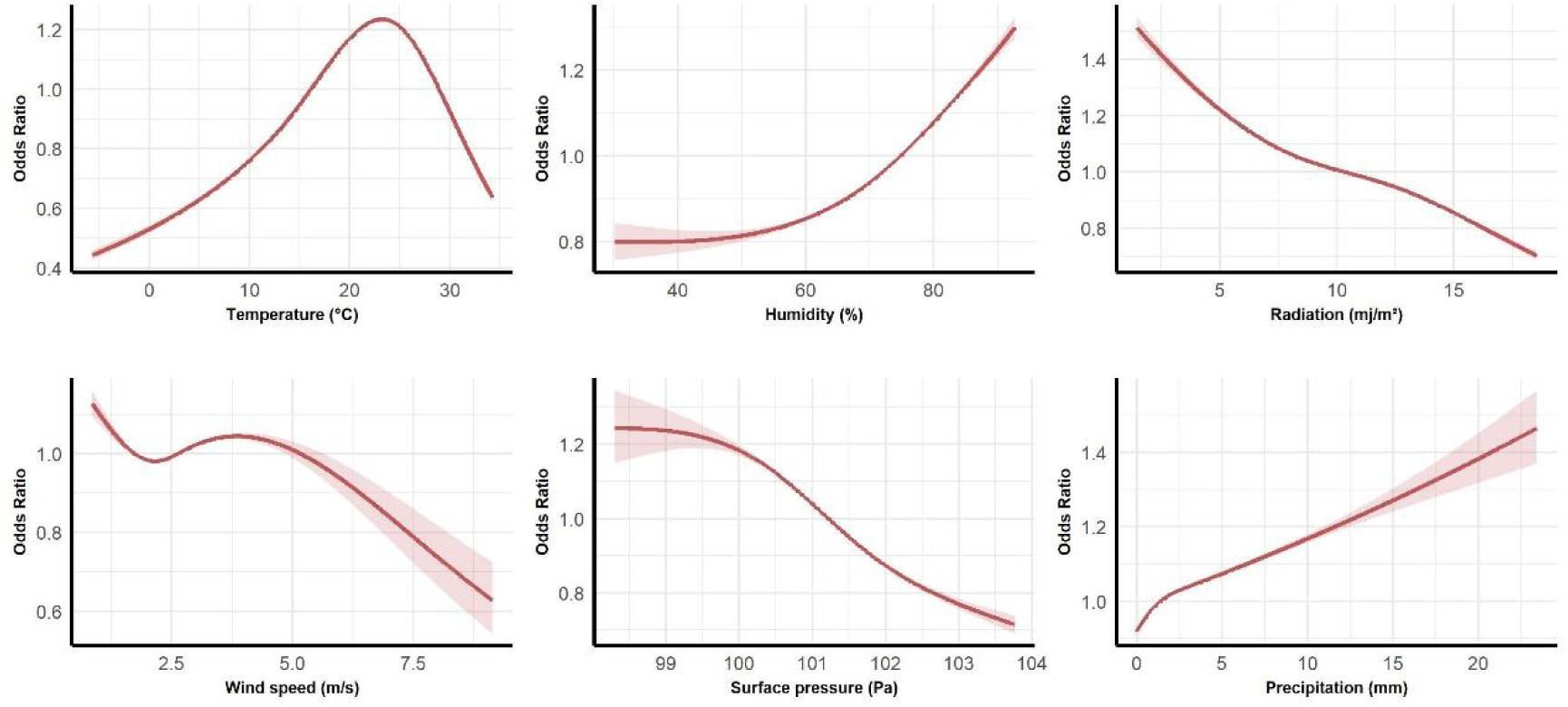
Exposure-response curves showing associations between 10-d moving average (lag010) exposures to six meteorological factors and risk of HFMD in Jiangsu, China, 2009-2023.

Figure 2 displays the exposure-response curves across the exposure continuum for the six meteorological factors over a 10-d moving average (lag010) using natural cubic spline models, which are adjusted for public holidays and variations in HFMD susceptibility. A consistent increasing trend is observed, with higher levels of humidity and precipitation associated with elevated HFMD risk, while an inverse relationship is found for radiation, wind speed, and surface pressure. For temperature, the curve follows a non-linear pattern, with HFMD risk initially increasing as the temperature rises to approximately 24 ℃, after which the risk begins to decline, indicating a potential threshold or optimal temperature range for HFMD risk.

### 3.4 Extreme meteorological factors and HFMD risk

To further explore the impact of extreme meteorological exposures on HFMD risk, we analyze the effects of exceeding the 85th, 90th, and 95th percentiles for each meteorological factor (Table 3). For temperature, each 1 ℃ increase above the 85th percentile threshold is associated with a 1.51% reduction in HFMD risk (OR = 0.9849, 95% CI: 0.9819, 0.9879). Similarly, each 1 ℃ increase above the 90th and 95th percentile corresponds to a 2.45% (OR = 0.9755, 95% CI: 0.9715, 0.9796) and a 5.04% (OR = 0.9496, 95% CI: 0.9426, 0.9566) decrease in risk, respectively. The analysis reveals excess radiation and surface pressure increase the risk of HFMD, with the positive association strengthening as higher thresholds are used to define excess magnitude. Conversely, excess temperature, humidity, wind speed, and precipitation reduce the risk of HFMD, with their negative associations becoming stronger as the thresholds increase.

**Table 3:** Associations between extreme meteorological exposures and HFMD risk at varying exposure thresholds in Jiangsu, China, 2009-2023.

| Variable | P85-OR (95% CI) | P90-OR (95% CI) | P95-OR (95% CI) |
| --- | --- | --- | --- |
| Temperature | 0.9849 (0.9819,0.9879) | 0.9755 (0.9715,0.9796) | 0.9496 (0.9426,0.9566) |
| Humidity | 0.9940 (0.9929,0.9952) | 0.9927 (0.9910,0.9944) | 0.9895 (0.9858,0.9932) |
| Wind speed | 0.9765 (0.9712,0.9818) | 0.9742 (0.9676,0.9808) | 0.9716 (0.9622,0.9811) |
| Radiation | 1.0168 (1.0127,1.0209) | 1.0315 (1.0247,1.0383) | 1.0769 (1.0613,1.0927) |
| Surface pressure | 1.0304 (1.0181,1.0428) | 1.0334 (1.0170,1.0500) | 1.0424 (1.0150,1.0707) |
| Precipitation | 0.9985 (0.9980,0.9989) | 0.9985 (0.9979,0.9990) | 0.9984 (0.9978,0.9991) |

### 3.5 Stratified analyses

The associations between meteorological exposures and HFMD, stratified by age, sex, and diagnostic type, for all single-lag models (lag 1 to lag 10) and moving average lag models (lag 01 to lag 010), are detailed in Table 4 and Tables S2-S7. Evidence of effect modification by age group is observed for temperature, humidity, and surface pressure ( *P* -interaction < 0.001). The association between temperature and HFMD is particularly pronounced among participants under 1 year of age. Male participants exhibit stronger associations between temperature and surface pressure with HFMD. In terms of diagnostic type, precipitation shows a more substantial effect among clinically confirmed cases compared to laboratory-confirmed cases.

**Table 4:**
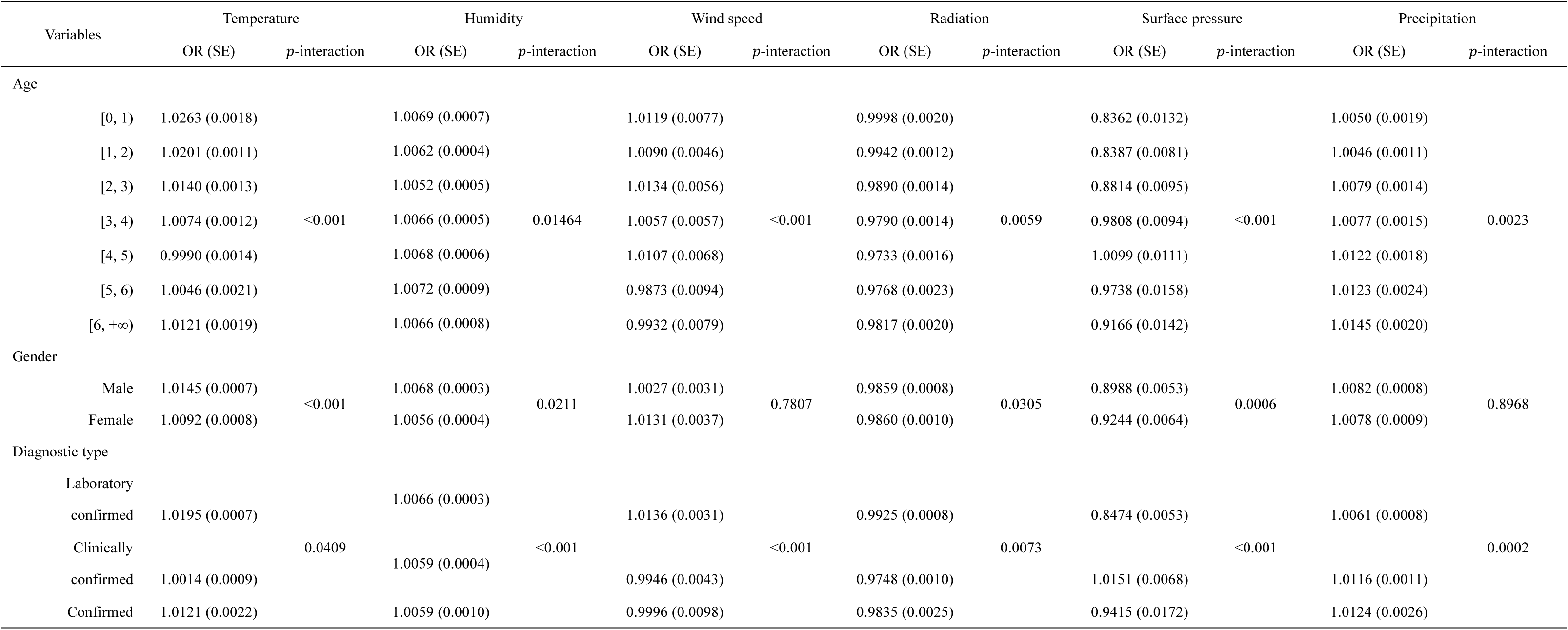
Associations of 10-d moving average (lag 010) exposures to meteorological factors with HFMD stratified by sex, age, diagnostic type in Jiangsu, China, 2009-2023.

### 3.6 Sensitivity analysis

Sensitivity analyses validate the robustness of our main findings. In the 2-meteorological factor models, the association between HFMD and individual meteorological factors remains robust after additional adjustment for the second factor, although some effect estimates are attenuated (Table S8). Additionally, DLNM models, adjusted for public holidays and HFMD susceptibility, display a similar trend to those in Figure 4 (Figure S4). Daytime exposure models also yield results consistent with our primary analysis (Figures S5 and S6). The overall trend remains consistent when restricting the analysis to 2009-2019 and 2020-2023, with relatively higher ORs for temperature, humidity, radiation, and precipitation during the 2020-2023 period (Tables S9 and S10).

## 4. Discussion

In this large case-crossover study, we find that exposure to higher temperature, humidity, wind speed, and precipitation is associated with an increased risk of HFMD in Jiangsu. We also observe a modest negative association between exposure to radiation, surface pressure, and HFMD. Notably, the association between HFMD and each meteorological factor usually remains significant in both the 2-meteorological model and the daytime exposure. The relationships of meteorological factors with HFMD vary by age, gender, and diagnostic type. As the thresholds for temperature, humidity, wind speed, and precipitation increase, the negative association between the excess magnitude and HFMD risk is strengthened, whereas the associations for radiation and surface pressure are reversed.

Our primary findings from the single-meteorological models corroborate previous time series city level studies. A study in California reports that HFMD incidence increases with average weekly temperature, while another investigation at the county level in Shaanxi Province identifies similar associations with rainfall, temperature, and relative humidity.^26,27^ In our current time-stratified case-crossover analysis, after controlling for potential confounders, we quantify the individual-level risk, yielding relatively conservative estimates of a 1.24%, 0.63%, and 0.80% higher HFMD risk at a moving lag of 10 days for each unit increase in temperature, humidity, and precipitation, respectively. To date, the mechanisms through which temperature and relative humidity alter the transmission risk of HFMD pathogens have been studied to some extent at the viral level.^11^ Laboratory studies confirm that, within certain temperature ranges, increases in both temperature and relative humidity elevate the infectiousness and viability of enteroviruses, thereby facilitating HFMD transmission within the community under favorable conditions.^28,29^ Notably, a threshold effect for temperature is evident in our analysis: the upward trend in HFMD risk reverses when average ambient temperatures exceed 24 ℃. Our findings echo similar results from Qingdao and Beijing, where HFMD incidence trends reverse at temperatures above 21 ℃ and 26 ℃.^30,31^ Early warning systems based on temperature threshold and high humidity could help predict and mitigate HFMD outbreaks, enabling timely interventions to reduce the disease burden. Previous studies suggest that the effect of average wind speed on HFMD risk is relatively weak;^31^ however, our findings indicate a positive association between wind speed and HFMD risk, in line with earlier research from Guangdong province.^33^

The negative association of surface pressure with HFMD aligns with findings from several prior studies.^33^ As a non-enveloped virus,^34^ the pathogens responsible for HFMD are particularly susceptible to environmental factors. Lower surface pressure, in particular, may facilitate increased water dispersion and promote the faster evaporation of aerosols, potentially enhancing viral transmission.^35^ To our knowledge, few studies have investigated the relationship between radiation and HFMD risk. This study quantifies a negative correlation between radiation and HFMD risk. Ultraviolet (UV) light, a key radiation component, has a well-documented germicidal effect, potentially reducing pathogen survival time in the environment and lowering HFMD transmission risk.^14,36^ It is worth noting that, due to the intricate interactions among various meteorological factors,^37,38^ isolating the independent effect of each factor and estimating the cumulative health impacts of multiple meteorological influences present substantial challenges. Further research is needed to address these complexities.

As the climate crisis intensifies, the frequency of extreme weather events has markedly increased.^39,40^ Previous studies have shown that extreme temperature events of varying intensity and duration impact mortality rates associated with myocardial infarction.^19^ However, the relationship between extreme weather and infectious diseases has received comparatively little attention.^41^ Our study examines the impact of extreme meteorological factors exceeding specific thresholds on HFMD risk. Notably, not all extreme weather conditions act as risk factors. For instance, the risk of HFMD decreases as temperature and wind speed exceed higher thresholds, whereas an increased risk is associated with radiation and surface pressure. Furthermore, the findings related to radiation diverge from those conclusions drawn in single-meteorological factor analyses, suggesting the necessity to distinguish between extreme effects and average effects.^42^ This distinction is pivotal for accurately identifying the highest risk and optimizing prevention and control strategies.^43^

Our stratified analyses suggest that the associations between temperature and HFMD risk are more potent in males and infants (< 1 year), consistent with previous findings.^18^ From a physiological perspective, infants possess immature immune and thermoregulatory systems, making them more susceptible to temperature fluctuations and less resistant to common environmental pathogens.^44^ Additionally, research indicates that males may exhibit heightened sensitivity to environmental factors compared to females, potentially due to gender differences in the frequency and duration of outdoor exposure.^18,45^ In contrast, the effect of precipitation on HFMD risk appears to be more pronounced in school-age children (> 6 years). Precipitation typically reduces the outdoor activity frequency in preschool-aged children, thereby limiting their exposure to environmental risk factors and lowering their infection risk.^46^

Our study possesses several strengths. First, the time-stratified case-crossover design provided individual-level evidence linking short-term meteorological exposure to the onset of HFMD symptoms, offering a unique opportunity to control for non-time-varying individual confounders, long-term temporal trends, and seasonality in the analysis. Second, the enhanced geographical resolution used in our exposure assessment reduced measurement error in exposure, which is often inevitable in previous epidemiological studies due to the unavailability of residential address data when deriving meteorological exposure from fixed monitoring stations.

Some limitations of our study warrant consideration. First, despite implementing rigorous quality control measures to identify HFMD cases, the large sample sizes may still be subject to uncertainties and underreporting within the monitoring data.^47^ However, sensitivity analyses, which restricted the sample to HFMD onset cases confirmed through reliable diagnostic criteria, demonstrated robust findings. Additionally, while meteorological data are derived from a grid dataset that accounts for outdoor exposure during the day, they do not capture the impact of indoor environmental factors, such as air conditioning or humidifiers. These indoor factors could significantly alter actual meteorological exposure levels,^48^ potentially affecting the interpretation of the study results.

In conclusion, our large-scale individual-level study employing a case-crossover design found that short-term exposure to ambient temperature, humidity, wind speed, and precipitation was associated with an elevated risk of HFMD. In contrast, exposure to radiation and surface pressure showed a modest negative association with HFMD risk. Gaining a deeper understanding of the meteorological drivers influencing HFMD virus transmission will help design early personalized interventions at individual and community levels and devise long-term prevention and control strategies targeting seasonal HFMD outbreaks.

## Data Availability

The data used in this study are not publicly available. Access to the data requires approval from the Jiangsu Provincial Center for Disease Control and Prevention in eastern China. For inquiries, please contact the corresponding author, Liguo Zhu.

## Declaration

### Ethics approval and consent to participate

The study is approved by the Xi’an Jiaotong-Liverpool University Research Ethics Committee (0010000089620241213145739).

### Consent for publication

Not applicable.

### Authors’ contributions

L.Z., C.L., and Y.W. contributed to the research design, conceptualization, and writing (review and editing). Y.T. prepared the initial manuscript draft and was responsible for coding and visualization. Y.T., H.Z., X.W., and K.W. processed the data and revised the initial draft. L.Z., H.J., and W.L. provided and managed the original data. L.B. and C.W. commented on and revised manuscript drafts. All authors read and approved the final manuscript.

## Acknowledgements

We thank the editors and anonymous reviewers for their helpful remarks.

## A. Appendix

**Table A.1:** Descriptions of six meteorological factors.

| Covariate | Description |
| --- | --- |
| Temperature (°C) | The air temperature at 2 meters above the surface of land, sea, or inland waters. |
| Humidity (%) | Calculated using the Clausius-Clapeyron equation based on air temperature and dew point temperature data. |
| Wind speed (m/s) | The square root of the sum of the squares of the northward and eastward components of the 10-meter wind vector. |
| Radiation (mj/m<sup>2</sup>) | The total amount of radiation reaching the Earth's surface, excluding the portion reflected back by the surface. |
| Surface pressure (kPa) | The atmospheric pressure is exerted at the Earth's surface, including land, sea, and inland waters. |
| Precipitation (mm) | The total accumulation of liquid and frozen water, including rain and snow, falls to the Earth's surface. |

