## Supplementary material for "Short-Term Effect of Ambient Meteorological Factors on Hand-Foot-Mouth Disease: An Individual-Level Case-Crossover Study in Jiangsu, China"

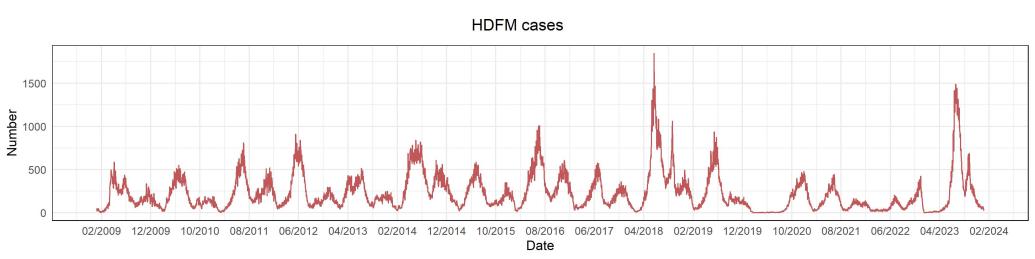

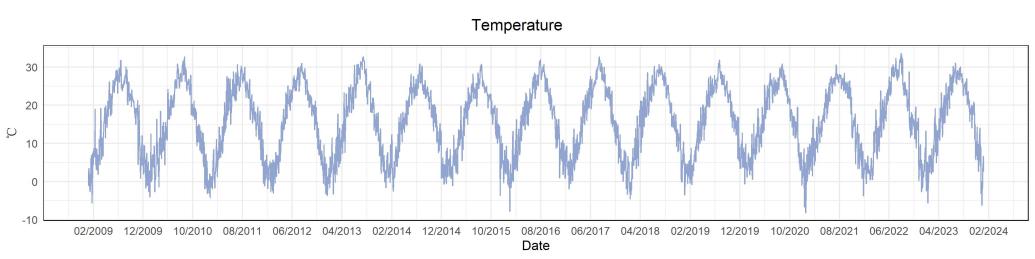

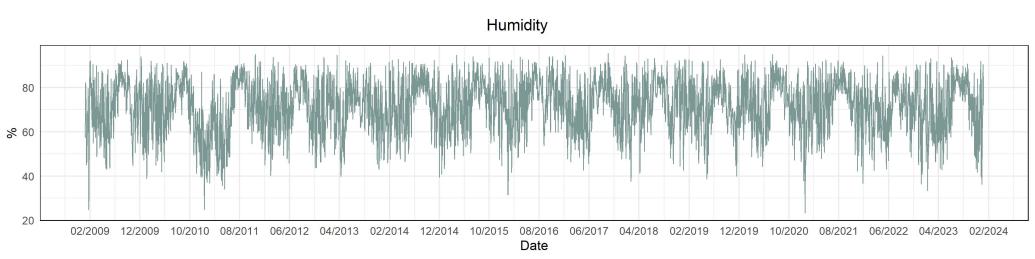

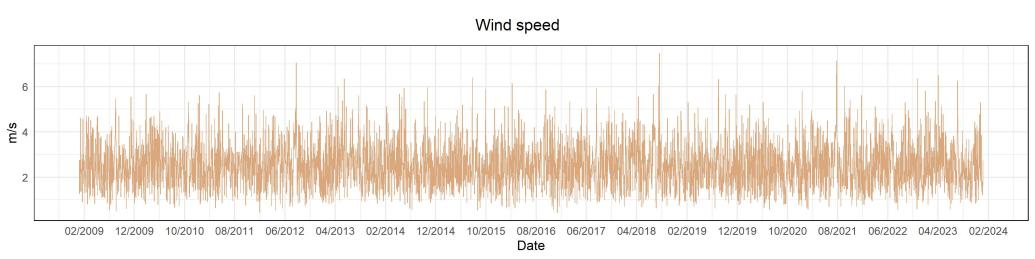

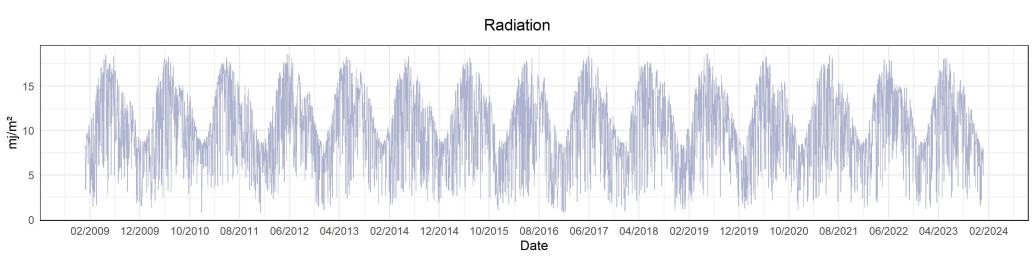

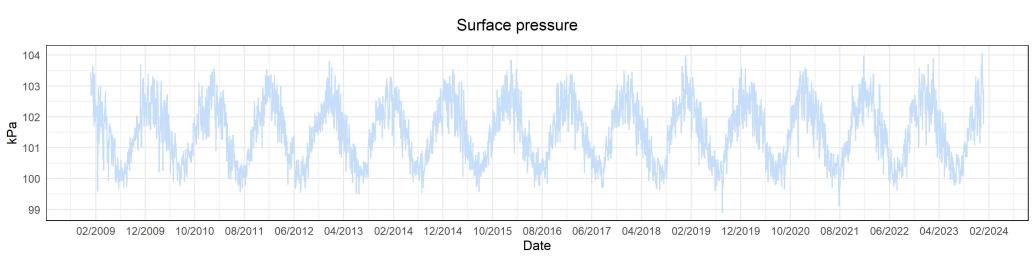

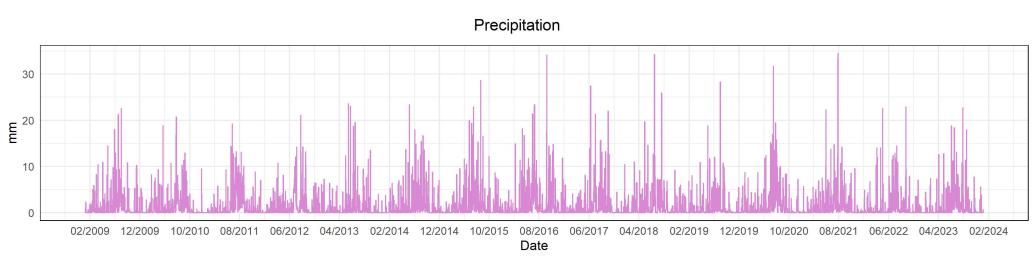

**Figure S1. Time-series plots for daily HMFD cases and meteorological factors in Jiangsu,** **China, 2009-2023.**

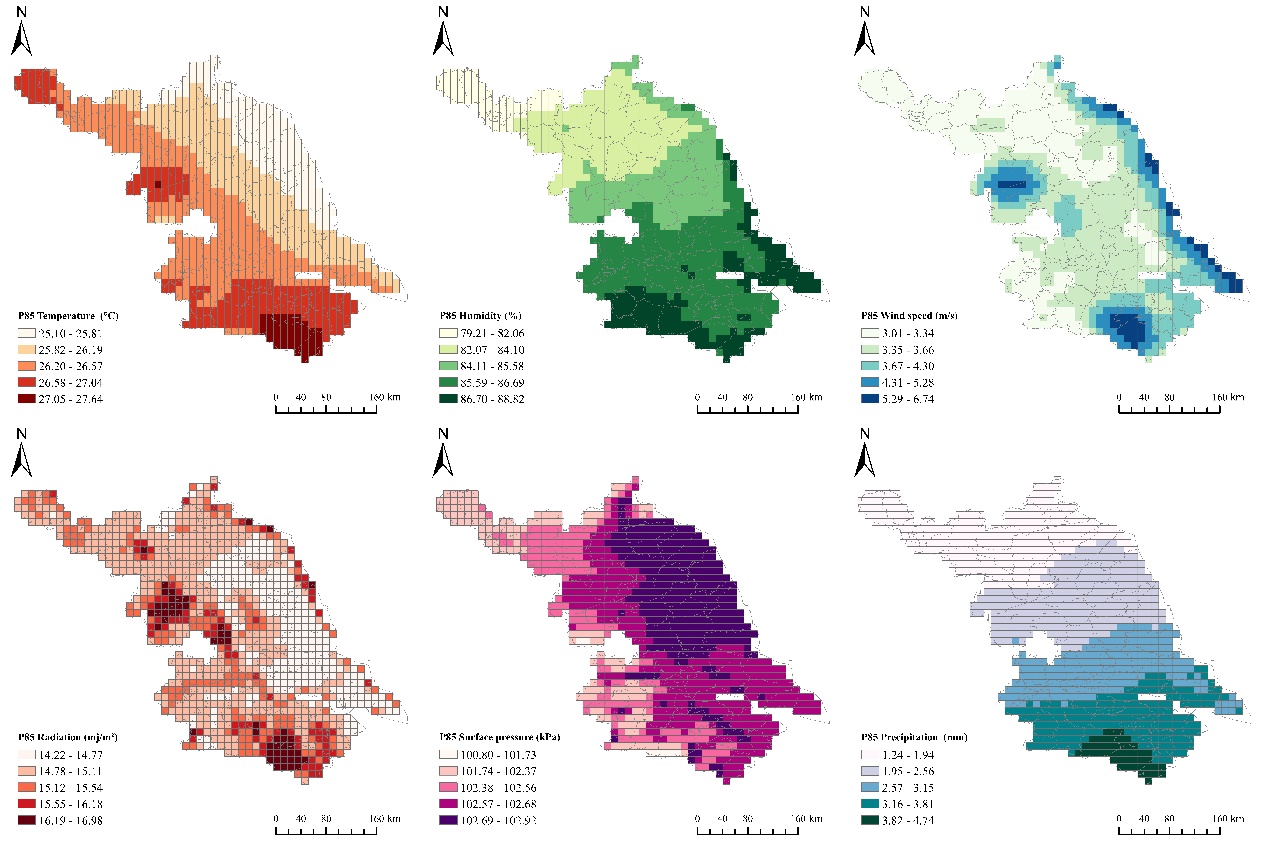

**Figure S2. Spatial distribution of meteorological factors 85th percentile thresholds in Jiangsu, China, 2009-2023.**

**Figure S3. Spatial distribution of meteorological factors 95th percentile thresholds in Jiangsu, China, 2009-2023.**

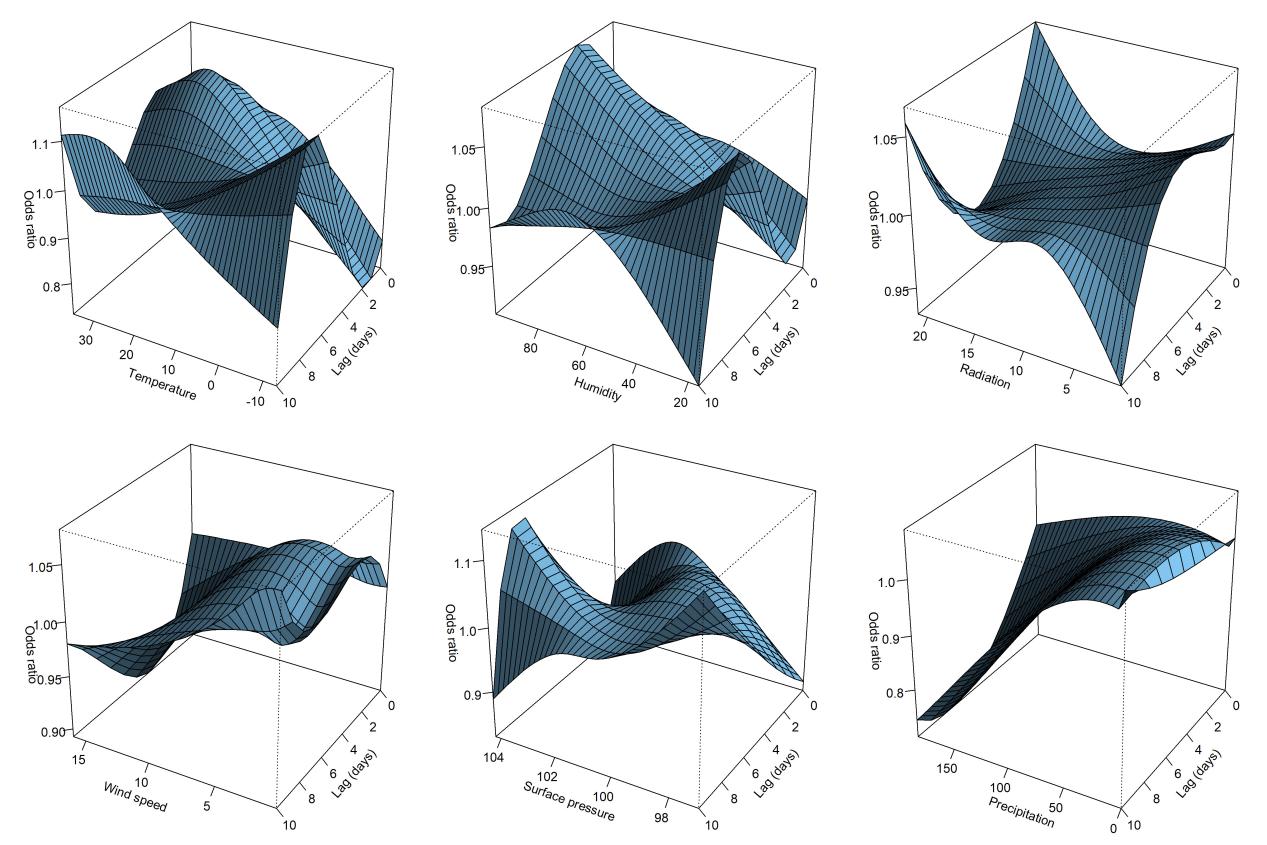

**Figure S4. Lag−response curves of meteorological factors and HFMD based on the DLNM model for Jiangsu, China from 2009 to 2023.**

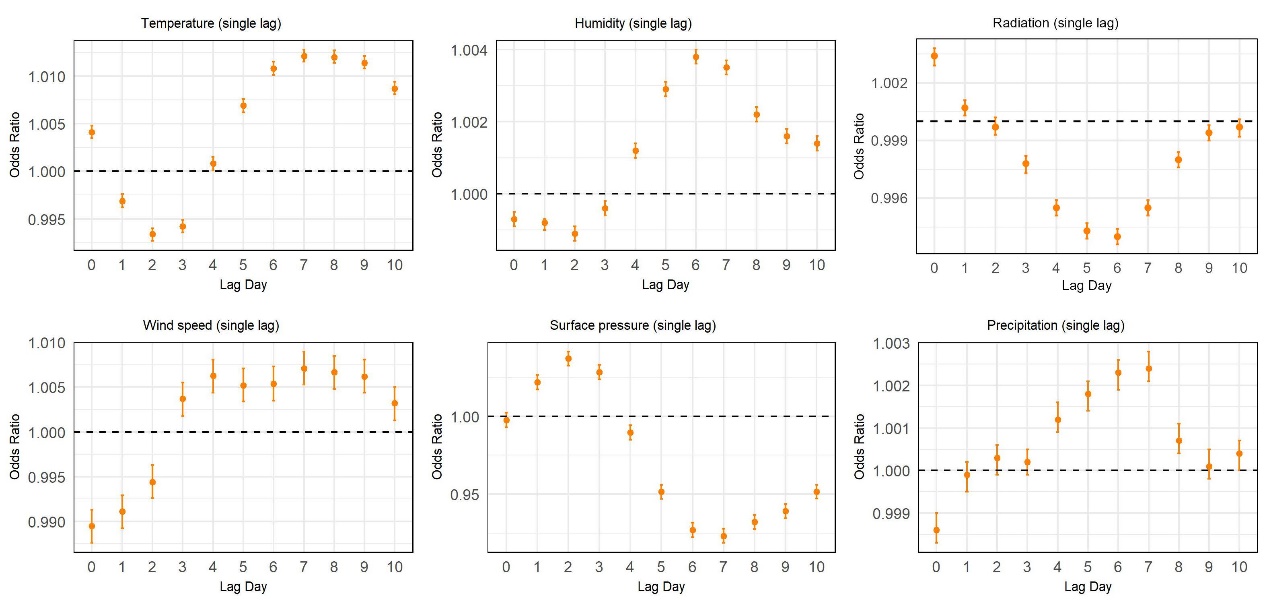

**Figure S5. Associations between acute daytime exposures to six meteorological factors and HFMD at different single lag days in Jiangsu, China, 2009-2023.**

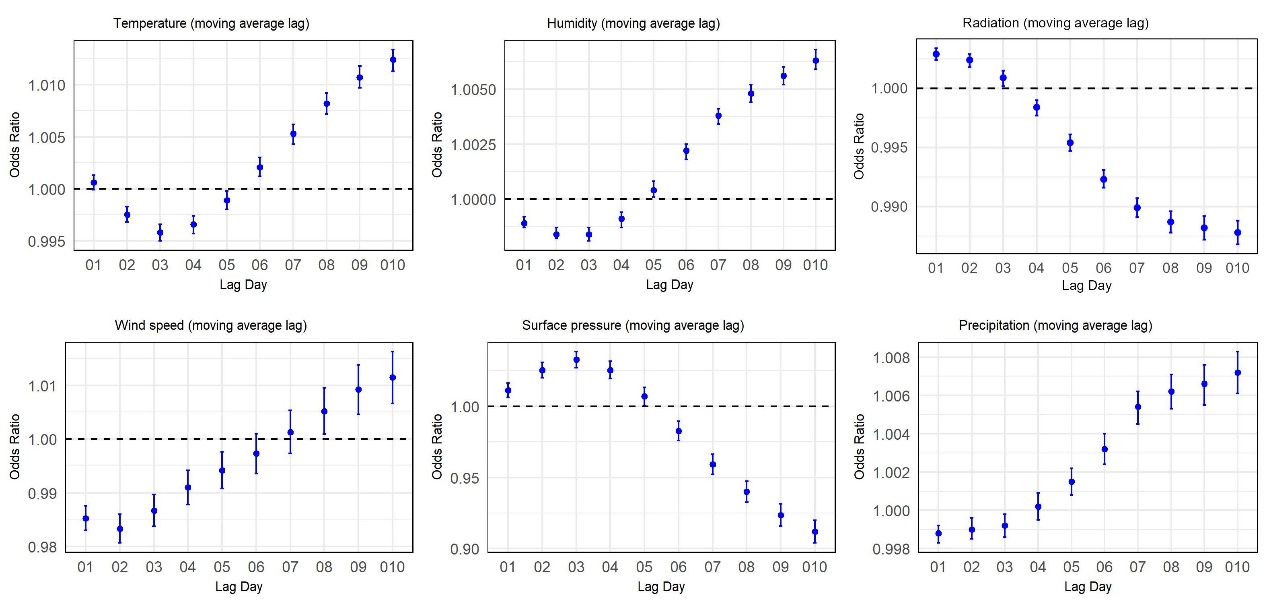

**Figure S6. Associations between acute daytime exposures to six meteorological factors and HFMD at different moving average lag days in Jiangsu, China, 2009-2023.**

**Table S1. Spearman correlations between six meteorological exposure factors for Jiangsu, China, 2009-2023.**

|  | Temperature | Humidity | Wind speed | Radiation | Surface pressure | Precipitation |
| --- | --- | --- | --- | --- | --- | --- |
| Temperature | 1.0000 | - | - | - | - | - |
| Humidity | 0.3588 | 1.0000 | - | - | - | - |
| Wind speed | 0.0382 | -0.0470 | 1.0000 | - | - | - |
| Radiation | 0.4195 | -0.4354 | -0.0034 | 1.0000 | - | - |
| Surface pressure | -0.8796 | -0.3892 | -0.0112 | -0.3295 | 1.0000 | - |
| Precipitation | 0.3154 | 0.6862 | 0.0967 | -0.4169 | -0.3825 | 1.0000 |

**Table S2 . OR (SD) of acute temperature exposure and HDFM at different single lag days and moving average lag days in Jiangsu, China, 2009-2023.**

|  | Age | | | | | | | Gender | | Diagnostic type | | |
| --- | --- | --- | --- | --- | --- | --- | --- | --- | --- | --- | --- | --- |
|  | ＜1 | 1-＜2 | 2-＜3 | 3-＜4 | 4-＜5 | 5-＜6 | ≥6 | Male | Female | Laboratory confirmed | Clinically confirmed | Confirmed |
| lag 1 | 1.0024(0.0011) | 0.9982(0.0007) | 0.9961(0.0008) | 0.9952(0.0008) | 0.9924(0.0009) | 0.9953(0.0013) | 0.9990(0.0012) | 0.9978(0.0004) | 0.9949(0.0005) | 0.9991(0.0004) | 0.9926(0.0006) | 0.9990(0.0015) |
| lag 2 | 1.0000(0.0011) | 0.9963(0.0007) | 0.9922(0.0008) | 0.9912(0.0008) | 0.9871(0.0009) | 0.9898(0.0013) | 0.9961(0.0012) | 0.9943(0.0004) | 0.9914(0.0005) | 0.9961(0.0004) | 0.9881(0.0006) | 0.9961(0.0015) |
| lag 3 | 1.0038(0.0011) | 0.9995(0.0007) | 0.9939(0.0008) | 0.9913(0.0008) | 0.9855(0.0009) | 0.9902(0.0013) | 0.9963(0.0012) | 0.9959(0.0004) | 0.9923(0.0005) | 0.9982(0.0004) | 0.9882(0.0006) | 0.9963(0.0015) |
| lag 4 | 1.0112(0.0011) | 1.0066(0.0007) | 1.0010(0.0008) | 0.9969(0.0008) | 0.9912(0.0009) | 0.9964(0.0013) | 1.0005(0.0012) | 1.0022(0.0004) | 0.9986(0.0005) | 1.0050(0.0004) | 0.9942(0.0006) | 1.0005(0.0015) |
| lag 5 | 1.0149(0.0011) | 1.0130(0.0007) | 1.0081(0.0008) | 1.0034(0.0008) | 0.9988(0.0009) | 1.0018(0.0013) | 1.0051(0.0012) | 1.0083(0.0004) | 1.0050(0.0005) | 1.0110(0.0004) | 1.0008(0.0006) | 1.0051(0.0014) |
| lag 6 | 1.0172(0.0011) | 1.0152(0.0007) | 1.0116(0.0008) | 1.0087(0.0008) | 1.0049(0.0009) | 1.0056(0.0013) | 1.0098(0.0012) | 1.0116(0.0004) | 1.0098(0.0005) | 1.0139(0.0004) | 1.0062(0.0006) | 1.0098(0.0014) |
| lag 7 | 1.0164(0.0011) | 1.0158(0.0007) | 1.0133(0.0008) | 1.0106(0.0008) | 1.0074(0.0009) | 1.0086(0.0013) | 1.0114(0.0012) | 1.0126(0.0004) | 1.0119(0.0005) | 1.0146(0.0004) | 1.0088(0.0006) | 1.0114(0.0014) |
| lag 8 | 1.0152(0.0011) | 1.0154(0.0007) | 1.0146(0.0008) | 1.0113(0.0008) | 1.0079(0.0009) | 1.0095(0.0013) | 1.0108(0.0012) | 1.0129(0.0004) | 1.0122(0.0005) | 1.0151(0.0004) | 1.0090(0.0006) | 1.0108(0.0014) |
| lag 9 | 1.0136(0.0011) | 1.0135(0.0007) | 1.0140(0.0008) | 1.0106(0.0008) | 1.0090(0.0009) | 1.0094(0.0013) | 1.0101(0.0012) | 1.0121(0.0004) | 1.0114(0.0005) | 1.0142(0.0004) | 1.0084(0.0005) | 1.0101(0.0014) |
| lag 10 | 1.0107(0.0011) | 1.0096(0.0007) | 1.0105(0.0008) | 1.0081(0.0008) | 1.0082(0.0009) | 1.0084(0.0013) | 1.0082(0.0012) | 1.0093(0.0004) | 1.0090(0.0005) | 1.0109(0.0004) | 1.0066(0.0005) | 1.0082(0.0014) |
| lag 01 | 1.0069(0.0012) | 1.0012(0.0007) | 1.0006(0.0009) | 0.9993(0.0008) | 0.9959(0.0010) | 0.9990(0.0014) | 1.0024(0.0013) | 1.0018(0.0005) | 0.9985(0.0006) | 1.0031(0.0005) | 0.9962(0.0006) | 1.0024(0.0016) |
| lag 02 | 1.0052(0.0013) | 0.9993(0.0008) | 0.9971(0.0009) | 0.9957(0.0009) | 0.9913(0.0010) | 0.9949(0.0015) | 1.0001(0.0014) | 0.9989(0.0005) | 0.9952(0.0006) | 1.0007(0.0005) | 0.9920(0.0006) | 1.0001(0.0017) |
| lag 03 | 1.0057(0.0014) | 0.9993(0.0008) | 0.9954(0.0010) | 0.9933(0.0009) | 0.9875(0.0011) | 0.9921(0.0016) | 0.9988(0.0015) | 0.9976(0.0005) | 0.9932(0.0007) | 0.9999(0.0005) | 0.9890(0.0007) | 0.9988(0.0018) |
| lag 04 | 1.0086(0.0014) | 1.0014(0.0009) | 0.9963(0.0010) | 0.9931(0.0010) | 0.9861(0.0012) | 0.9919(0.0017) | 0.9990(0.0015) | 0.9986(0.0006) | 0.9936(0.0007) | 1.0015(0.0006) | 0.9885(0.0007) | 0.9990(0.0018) |
| lag 05 | 1.0121(0.0015) | 1.0050(0.0009) | 0.9989(0.0011) | 0.9947(0.0010) | 0.9870(0.0012) | 0.9931(0.0018) | 1.0006(0.0016) | 1.0011(0.0006) | 0.9956(0.0007) | 1.0045(0.0006) | 0.9897(0.0008) | 1.0006(0.0019) |
| lag 06 | 1.0161(0.0016) | 1.0089(0.0009) | 1.0022(0.0011) | 0.9975(0.0011) | 0.9893(0.0013) | 0.9952(0.0018) | 1.0033(0.0016) | 1.0043(0.0006) | 0.9986(0.0007) | 1.0081(0.0006) | 0.9922(0.0008) | 1.0033(0.0020) |
| lag 07 | 1.0196(0.0016) | 1.0126(0.0010) | 1.0056(0.0011) | 1.0005(0.0011) | 0.9919(0.0013) | 0.9977(0.0019) | 1.0061(0.0017) | 1.0074(0.0006) | 1.0018(0.0008) | 1.0115(0.0006) | 0.9950(0.0008) | 1.0061(0.0021) |
| lag 08 | 1.0226(0.0017) | 1.0160(0.0010) | 1.0090(0.0012) | 1.0033(0.0012) | 0.9943(0.0014) | 1.0003(0.0020) | 1.0086(0.0018) | 1.0103(0.0007) | 1.0048(0.0008) | 1.0148(0.0007) | 0.9976(0.0008) | 1.0086(0.0021) |
| lag 09 | 1.0250(0.0017) | 1.0186(0.0011) | 1.0120(0.0012) | 1.0057(0.0012) | 0.9968(0.0014) | 1.0027(0.0020) | 1.0107(0.0018) | 1.0129(0.0007) | 1.0074(0.0008) | 1.0177(0.0007) | 0.9998(0.0009) | 1.0107(0.0022) |
| lag 010 | 1.0263(0.0018) | 1.0201(0.0011) | 1.0140(0.0013) | 1.0074(0.0012) | 0.9990(0.0014) | 1.0046(0.0021) | 1.0121(0.0019) | 1.0145(0.0007) | 1.0092(0.0008) | 1.0195(0.0007) | 1.0014(0.0009) | 1.0121(0.0022) |

**Table S3 . OR (SD) of acute humidity exposure and HDFM at different single lag days and moving average lag days in Jiangsu, China, 2009-2023.**

|  | Age | | | | | | | Gender | | Diagnostic type | | |
| --- | --- | --- | --- | --- | --- | --- | --- | --- | --- | --- | --- | --- |
|  | ＜1 | 1-＜2 | 2-＜3 | 3-＜4 | 4-＜5 | 5-＜6 | ≥6 | Male | Female | Laboratory confirmed | Clinically confirmed | Confirmed |
| lag 1 | 1.0001(0.0003) | 0.9987(0.0002) | 0.9986(0.0002) | 0.9990(0.0002) | 0.9989(0.0003) | 0.9986(0.0004) | 1.0000(0.0004) | 0.9991(0.0001) | 0.9989(0.0002) | 0.9990(0.0001) | 0.9988(0.0002) | 1.0000(0.0005) |
| lag 2 | 1.0005(0.0003) | 0.9988(0.0002) | 0.9984(0.0002) | 0.9987(0.0002) | 0.9984(0.0003) | 0.9986(0.0004) | 0.9992(0.0004) | 0.9990(0.0001) | 0.9986(0.0002) | 0.9990(0.0001) | 0.9985(0.0002) | 0.9991(0.0005) |
| lag 3 | 1.0013(0.0003) | 1.0001(0.0002) | 0.9993(0.0002) | 0.9997(0.0002) | 0.9989(0.0003) | 1.0001(0.0004) | 0.9995(0.0004) | 1.0000(0.0001) | 0.9995(0.0002) | 1.0001(0.0001) | 0.9994(0.0002) | 0.9993(0.0004) |
| lag 4 | 1.0026(0.0003) | 1.0017(0.0002) | 1.0016(0.0002) | 1.0014(0.0002) | 1.0009(0.0003) | 1.0022(0.0004) | 1.0007(0.0004) | 1.0018(0.0001) | 1.0011(0.0002) | 1.0019(0.0001) | 1.0012(0.0002) | 1.0001(0.0004) |
| lag 5 | 1.0035(0.0003) | 1.0031(0.0002) | 1.0033(0.0002) | 1.0035(0.0002) | 1.0033(0.0003) | 1.0033(0.0004) | 1.0027(0.0004) | 1.0035(0.0001) | 1.0028(0.0002) | 1.0033(0.0001) | 1.0034(0.0002) | 1.0021(0.0005) |
| lag 6 | 1.0032(0.0003) | 1.0039(0.0002) | 1.0036(0.0002) | 1.0047(0.0002) | 1.0049(0.0003) | 1.0041(0.0004) | 1.0039(0.0004) | 1.0042(0.0001) | 1.0039(0.0002) | 1.0038(0.0001) | 1.0047(0.0002) | 1.0030(0.0005) |
| lag 7 | 1.0023(0.0003) | 1.0033(0.0002) | 1.0033(0.0002) | 1.0043(0.0002) | 1.0045(0.0003) | 1.0042(0.0004) | 1.0034(0.0004) | 1.0036(0.0001) | 1.0037(0.0002) | 1.0034(0.0001) | 1.0041(0.0002) | 1.0028(0.0004) |
| lag 8 | 1.0017(0.0003) | 1.0023(0.0002) | 1.0023(0.0002) | 1.0022(0.0002) | 1.0027(0.0003) | 1.0024(0.0004) | 1.0018(0.0004) | 1.0022(0.0001) | 1.0023(0.0002) | 1.0025(0.0001) | 1.0019(0.0002) | 1.0025(0.0004) |
| lag 9 | 1.0011(0.0003) | 1.0020(0.0002) | 1.0015(0.0002) | 1.0012(0.0002) | 1.0019(0.0003) | 1.0016(0.0004) | 1.0014(0.0004) | 1.0016(0.0001) | 1.0015(0.0002) | 1.0019(0.0001) | 1.0010(0.0002) | 1.0023(0.0004) |
| lag 10 | 1.0008(0.0003) | 1.0014(0.0002) | 1.0014(0.0002) | 1.0011(0.0002) | 1.0015(0.0003) | 1.0014(0.0004) | 1.0019(0.0004) | 1.0012(0.0001) | 1.0015(0.0002) | 1.0015(0.0001) | 1.0010(0.0002) | 1.0019(0.0004) |
| lag 01 | 0.9996(0.0004) | 0.9985(0.0002) | 0.9983(0.0003) | 0.9988(0.0003) | 0.9984(0.0003) | 0.9985(0.0005) | 1.0001(0.0004) | 0.9988(0.0002) | 0.9986(0.0002) | 0.9987(0.0002) | 0.9985(0.0002) | 1.0002(0.0005) |
| lag 02 | 1.0000(0.0004) | 0.9981(0.0003) | 0.9977(0.0003) | 0.9983(0.0003) | 0.9978(0.0004) | 0.9979(0.0005) | 0.9996(0.0005) | 0.9985(0.0002) | 0.9981(0.0002) | 0.9984(0.0002) | 0.9979(0.0002) | 0.9996(0.0006) |
| lag 03 | 1.0006(0.0005) | 0.9984(0.0003) | 0.9976(0.0003) | 0.9983(0.0003) | 0.9974(0.0004) | 0.9982(0.0006) | 0.9994(0.0005) | 0.9986(0.0002) | 0.9980(0.0002) | 0.9986(0.0002) | 0.9978(0.0002) | 0.9993(0.0006) |
| lag 04 | 1.0017(0.0005) | 0.9993(0.0003) | 0.9985(0.0004) | 0.9990(0.0004) | 0.9980(0.0004) | 0.9993(0.0006) | 0.9998(0.0006) | 0.9995(0.0002) | 0.9987(0.0002) | 0.9996(0.0002) | 0.9985(0.0003) | 0.9994(0.0007) |
| lag 05 | 1.0031(0.0005) | 1.0006(0.0003) | 1.0000(0.0004) | 1.0006(0.0004) | 0.9996(0.0004) | 1.0009(0.0006) | 1.0010(0.0006) | 1.0011(0.0002) | 1.0000(0.0003) | 1.0010(0.0002) | 1.0001(0.0003) | 1.0003(0.0007) |
| lag 06 | 1.0043(0.0006) | 1.0022(0.0003) | 1.0015(0.0004) | 1.0026(0.0004) | 1.0018(0.0005) | 1.0027(0.0007) | 1.0027(0.0006) | 1.0029(0.0002) | 1.0016(0.0003) | 1.0026(0.0002) | 1.0021(0.0003) | 1.0016(0.0008) |
| lag 07 | 1.0053(0.0006) | 1.0036(0.0004) | 1.0029(0.0004) | 1.0045(0.0004) | 1.0038(0.0005) | 1.0046(0.0007) | 1.0042(0.0007) | 1.0044(0.0002) | 1.0032(0.0003) | 1.0040(0.0002) | 1.0039(0.0003) | 1.0029(0.0008) |
| lag 08 | 1.0060(0.0006) | 1.0046(0.0004) | 1.0039(0.0005) | 1.0055(0.0005) | 1.0051(0.0005) | 1.0057(0.0008) | 1.0050(0.0007) | 1.0054(0.0003) | 1.0042(0.0003) | 1.0051(0.0003) | 1.0048(0.0003) | 1.0040(0.0009) |
| lag 09 | 1.0065(0.0007) | 1.0055(0.0004) | 1.0046(0.0005) | 1.0061(0.0005) | 1.0060(0.0006) | 1.0065(0.0008) | 1.0057(0.0008) | 1.0062(0.0003) | 1.0049(0.0003) | 1.0059(0.0003) | 1.0053(0.0004) | 1.0050(0.0009) |
| lag 010 | 1.0069(0.0007) | 1.0062(0.0004) | 1.0052(0.0005) | 1.0066(0.0005) | 1.0068(0.0006) | 1.0072(0.0009) | 1.0066(0.0008) | 1.0068(0.0003) | 1.0056(0.0004) | 1.0066(0.0003) | 1.0059(0.0004) | 1.0059(0.0010) |

**Table S4 . OR (SD) of acute wind speed exposure and HDFM at different single lag days and moving average lag days in Jiangsu, China, 2009-2023.**

|  | Age | | | | | | | Gender | | Diagnostic type | | |
| --- | --- | --- | --- | --- | --- | --- | --- | --- | --- | --- | --- | --- |
|  | ＜1 | 1-＜2 | 2-＜3 | 3-＜4 | 4-＜5 | 5-＜6 | ≥6 | Male | Female | Laboratory confirmed | Clinically confirmed | Confirmed |
| lag 1 | 0.9963(0.0031) | 0.9998(0.0018) | 0.9935(0.0022) | 0.9845(0.0022) | 0.9840(0.0025) | 0.9856(0.0035) | 0.9968(0.0031) | 0.9910(0.0012) | 0.9940(0.0015) | 0.9969(0.0012) | 0.9862(0.0016) | 1.0009(0.0039) |
| lag 2 | 1.0013(0.0031) | 1.0043(0.0018) | 0.9993(0.0022) | 0.9893(0.0022) | 0.9911(0.0025) | 0.9922(0.0035) | 0.9923(0.0031) | 0.9955(0.0012) | 0.9986(0.0015) | 1.0012(0.0012) | 0.9885(0.0016) | 0.9992(0.0039) |
| lag 3 | 1.0078(0.0031) | 1.0063(0.0018) | 1.0059(0.0022) | 1.0012(0.0022) | 1.0044(0.0025) | 1.0004(0.0035) | 1.0047(0.0031) | 1.0036(0.0012) | 1.0061(0.0015) | 1.0066(0.0012) | 1.0014(0.0016) | 1.0034(0.0039) |
| lag 4 | 1.0057(0.0031) | 1.0054(0.0018) | 1.0065(0.0022) | 1.0093(0.0022) | 1.0072(0.0025) | 1.0016(0.0035) | 1.0035(0.0032) | 1.0051(0.0012) | 1.0077(0.0015) | 1.0065(0.0012) | 1.0053(0.0016) | 1.0073(0.0039) |
| lag 5 | 1.0046(0.0031) | 1.0023(0.0018) | 1.0031(0.0022) | 1.0070(0.0022) | 1.0084(0.0026) | 1.0030(0.0036) | 1.0022(0.0032) | 1.0032(0.0012) | 1.0060(0.0015) | 1.0040(0.0012) | 1.0045(0.0016) | 1.0071(0.0039) |
| lag 6 | 1.0038(0.0031) | 1.0011(0.0018) | 1.0090(0.0022) | 1.0046(0.0022) | 1.0061(0.0026) | 1.0020(0.0036) | 0.9989(0.0032) | 1.0029(0.0012) | 1.0055(0.0015) | 1.0033(0.0012) | 1.0059(0.0016) | 0.9989(0.0039) |
| lag 7 | 1.0071(0.0031) | 1.0040(0.0018) | 1.0090(0.0022) | 1.0076(0.0022) | 1.0086(0.0025) | 1.0033(0.0036) | 0.9985(0.0032) | 1.0065(0.0012) | 1.0053(0.0015) | 1.0056(0.0012) | 1.0078(0.0016) | 0.9971(0.0040) |
| lag 8 | 0.9994(0.0031) | 1.0009(0.0018) | 1.0042(0.0022) | 1.0135(0.0022) | 1.0121(0.0025) | 1.0054(0.0036) | 0.9948(0.0032) | 1.0059(0.0012) | 1.0039(0.0015) | 1.0029(0.0012) | 1.0110(0.0016) | 0.9894(0.0040) |
| lag 9 | 1.0030(0.0031) | 1.0004(0.0018) | 1.0026(0.0022) | 1.0069(0.0022) | 1.0090(0.0025) | 1.0039(0.0035) | 1.0027(0.0032) | 1.0031(0.0012) | 1.0051(0.0015) | 1.0034(0.0012) | 1.0059(0.0016) | 0.9965(0.0039) |
| lag 10 | 0.9999(0.0031) | 0.9999(0.0018) | 0.9993(0.0022) | 1.0026(0.0022) | 1.0016(0.0025) | 0.9998(0.0035) | 0.9987(0.0031) | 1.0001(0.0012) | 1.0008(0.0015) | 1.0015(0.0012) | 0.9984(0.0016) | 1.0012(0.0039) |
| lag 01 | 0.9912(0.0037) | 0.9935(0.0022) | 0.9881(0.0026) | 0.9764(0.0026) | 0.9769(0.0031) | 0.9771(0.0043) | 0.9940(0.0038) | 0.9847(0.0014) | 0.9878(0.0018) | 0.9917(0.0014) | 0.9734(0.0019) | 0.9991(0.0047) |
| lag 02 | 0.9932(0.0043) | 0.9971(0.0025) | 0.9892(0.0030) | 0.9723(0.0030) | 0.9737(0.0036) | 0.9749(0.0049) | 0.9900(0.0043) | 0.9837(0.0017) | 0.9885(0.0020) | 0.9936(0.0017) | 0.9689(0.0022) | 0.9987(0.0053) |
| lag 03 | 0.9983(0.0047) | 1.0010(0.0028) | 0.9935(0.0033) | 0.9750(0.0034) | 0.9781(0.0040) | 0.9769(0.0055) | 0.9935(0.0047) | 0.9871(0.0018) | 0.9929(0.0022) | 0.9979(0.0018) | 0.9718(0.0025) | 1.0008(0.0059) |
| lag 04 | 1.0015(0.0051) | 1.0039(0.0030) | 0.9974(0.0036) | 0.9813(0.0037) | 0.9831(0.0043) | 0.9789(0.0060) | 0.9957(0.0052) | 0.9905(0.0020) | 0.9975(0.0024) | 1.0016(0.0020) | 0.9759(0.0027) | 1.0047(0.0064) |
| lag 05 | 1.0039(0.0055) | 1.0050(0.0032) | 0.9991(0.0039) | 0.9857(0.0040) | 0.9882(0.0047) | 0.9810(0.0065) | 0.9970(0.0055) | 0.9925(0.0022) | 1.0008(0.0026) | 1.0036(0.0022) | 0.9790(0.0030) | 1.0082(0.0069) |
| lag 06 | 1.0059(0.0059) | 1.0054(0.0035) | 1.0039(0.0042) | 0.9884(0.0043) | 0.9917(0.0051) | 0.9822(0.0070) | 0.9965(0.0059) | 0.9941(0.0023) | 1.0037(0.0028) | 1.0053(0.0023) | 0.9824(0.0032) | 1.0075(0.0074) |
| lag 07 | 1.0098(0.0064) | 1.0076(0.0037) | 1.0089(0.0046) | 0.9922(0.0046) | 0.9963(0.0055) | 0.9835(0.0076) | 0.9957(0.0064) | 0.9975(0.0025) | 1.0066(0.0031) | 1.0083(0.0025) | 0.9863(0.0035) | 1.0061(0.0079) |
| lag 08 | 1.0098(0.0068) | 1.0084(0.0040) | 1.0115(0.0049) | 0.9998(0.0050) | 1.0035(0.0060) | 0.9859(0.0082) | 0.9929(0.0069) | 1.0007(0.0027) | 1.0091(0.0033) | 1.0102(0.0027) | 0.9924(0.0037) | 1.0008(0.0086) |
| lag 09 | 1.0117(0.0073) | 1.0088(0.0043) | 1.0135(0.0052) | 1.0040(0.0053) | 1.0094(0.0064) | 0.9878(0.0088) | 0.9941(0.0074) | 1.0025(0.0029) | 1.0123(0.0035) | 1.0124(0.0029) | 0.9959(0.0040) | 0.9989(0.0092) |
| lag 010 | 1.0119(0.0077) | 1.0090(0.0046) | 1.0134(0.0056) | 1.0057(0.0057) | 1.0107(0.0068) | 0.9873(0.0094) | 0.9932(0.0079) | 1.0027(0.0031) | 1.0131(0.0037) | 1.0136(0.0031) | 0.9946(0.0043) | 0.9996(0.0098) |

**Table S5 . OR (SD) of acute radiation exposure and HDFM at different single lag days and moving average lag days in Jiangsu, China, 2009-2023.**

|  | Age | | | | | | | Gender | | Diagnostic type | | |
| --- | --- | --- | --- | --- | --- | --- | --- | --- | --- | --- | --- | --- |
|  | ＜1 | 1-＜2 | 2-＜3 | 3-＜4 | 4-＜5 | 5-＜6 | ≥6 | Male | Female | Laboratory confirmed | Clinically confirmed | Confirmed |
| lag 1 | 1.0003(0.0008) | 1.0015(0.0005) | 1.0017(0.0006) | 1.0008(0.0006) | 1.0003(0.0007) | 1.0007(0.0009) | 0.9982(0.0009) | 1.0008(0.0003) | 1.0008(0.0004) | 1.0016(0.0003) | 1.0001(0.0004) | 0.9965(0.0011) |
| lag 2 | 0.9993(0.0008) | 1.0001(0.0005) | 0.9998(0.0006) | 0.9994(0.0006) | 0.9996(0.0007) | 0.9988(0.0010) | 0.9985(0.0009) | 0.9996(0.0003) | 0.9996(0.0004) | 1.0001(0.0003) | 0.9988(0.0004) | 0.9984(0.0011) |
| lag 3 | 0.9982(0.0008) | 0.9978(0.0005) | 0.9972(0.0006) | 0.9962(0.0006) | 0.9968(0.0007) | 0.9952(0.0009) | 0.9976(0.0008) | 0.9971(0.0003) | 0.9971(0.0004) | 0.9978(0.0003) | 0.9960(0.0004) | 0.9974(0.0011) |
| lag 4 | 0.9972(0.0008) | 0.9968(0.0005) | 0.9938(0.0006) | 0.9926(0.0006) | 0.9921(0.0007) | 0.9924(0.0009) | 0.9952(0.0008) | 0.9942(0.0003) | 0.9948(0.0004) | 0.9958(0.0003) | 0.9919(0.0004) | 0.9958(0.0011) |
| lag 5 | 0.9967(0.0008) | 0.9956(0.0005) | 0.9933(0.0006) | 0.9904(0.0006) | 0.9896(0.0007) | 0.9907(0.0009) | 0.9930(0.0009) | 0.9928(0.0003) | 0.9932(0.0004) | 0.9949(0.0003) | 0.9894(0.0004) | 0.9943(0.0011) |
| lag 6 | 0.9984(0.0008) | 0.9955(0.0005) | 0.9937(0.0006) | 0.9898(0.0006) | 0.9882(0.0007) | 0.9898(0.0009) | 0.9924(0.0008) | 0.9928(0.0003) | 0.9926(0.0004) | 0.9952(0.0003) | 0.9881(0.0004) | 0.9953(0.0011) |
| lag 7 | 1.0006(0.0008) | 0.9974(0.0005) | 0.9951(0.0006) | 0.9915(0.0006) | 0.9904(0.0007) | 0.9917(0.0009) | 0.9952(0.0008) | 0.9948(0.0003) | 0.9944(0.0004) | 0.9965(0.0003) | 0.9910(0.0004) | 0.9975(0.0011) |
| lag 8 | 1.0017(0.0008) | 0.9992(0.0005) | 0.9981(0.0006) | 0.9963(0.0006) | 0.9945(0.0007) | 0.9962(0.0009) | 0.9979(0.0008) | 0.9978(0.0003) | 0.9977(0.0004) | 0.9985(0.0003) | 0.9964(0.0004) | 0.9978(0.0011) |
| lag 9 | 1.0018(0.0008) | 1.0001(0.0005) | 1.0005(0.0006) | 0.9992(0.0006) | 0.9964(0.0007) | 0.9982(0.0009) | 0.9982(0.0008) | 0.9992(0.0003) | 0.9995(0.0004) | 1.0001(0.0003) | 0.9985(0.0004) | 0.9970(0.0011) |
| lag 10 | 1.0010(0.0008) | 1.0006(0.0005) | 1.0007(0.0006) | 0.9993(0.0006) | 0.9980(0.0007) | 0.9996(0.0009) | 0.9977(0.0008) | 1.0000(0.0003) | 0.9993(0.0004) | 1.0003(0.0003) | 0.9991(0.0004) | 0.9975(0.0011) |
| lag 01 | 1.0034(0.0010) | 1.0039(0.0006) | 1.0047(0.0007) | 1.0036(0.0007) | 1.0033(0.0008) | 1.0025(0.0011) | 0.9986(0.0010) | 1.0034(0.0004) | 1.0033(0.0005) | 1.0044(0.0004) | 1.0026(0.0005) | 0.9966(0.0013) |
| lag 02 | 1.0025(0.0011) | 1.0035(0.0007) | 1.0039(0.0008) | 1.0027(0.0008) | 1.0026(0.0009) | 1.0014(0.0013) | 0.9979(0.0011) | 1.0027(0.0004) | 1.0026(0.0005) | 1.0039(0.0004) | 1.0015(0.0005) | 0.9962(0.0014) |
| lag 03 | 1.0013(0.0013) | 1.0020(0.0007) | 1.0020(0.0009) | 1.0004(0.0009) | 1.0007(0.0010) | 0.9986(0.0014) | 0.9968(0.0013) | 1.0009(0.0005) | 1.0007(0.0006) | 1.0023(0.0005) | 0.9991(0.0006) | 0.9951(0.0016) |
| lag 04 | 0.9997(0.0014) | 1.0002(0.0008) | 0.9986(0.0010) | 0.9965(0.0010) | 0.9964(0.0011) | 0.9946(0.0015) | 0.9944(0.0014) | 0.9977(0.0005) | 0.9980(0.0006) | 1.0000(0.0005) | 0.9948(0.0006) | 0.9932(0.0017) |
| lag 05 | 0.9981(0.0015) | 0.9980(0.0009) | 0.9953(0.0010) | 0.9918(0.0010) | 0.9911(0.0012) | 0.9901(0.0017) | 0.9912(0.0015) | 0.9942(0.0006) | 0.9947(0.0007) | 0.9974(0.0006) | 0.9896(0.0007) | 0.9907(0.0018) |
| lag 06 | 0.9973(0.0016) | 0.9958(0.0009) | 0.9923(0.0011) | 0.9869(0.0011) | 0.9853(0.0013) | 0.9851(0.0018) | 0.9876(0.0016) | 0.9907(0.0006) | 0.9911(0.0007) | 0.9952(0.0006) | 0.9837(0.0007) | 0.9886(0.0020) |
| lag 07 | 0.9976(0.0017) | 0.9946(0.0010) | 0.9898(0.0012) | 0.9825(0.0012) | 0.9802(0.0014) | 0.9807(0.0019) | 0.9852(0.0017) | 0.9881(0.0006) | 0.9883(0.0008) | 0.9934(0.0006) | 0.9790(0.0008) | 0.9874(0.0021) |
| lag 08 | 0.9984(0.0018) | 0.9941(0.0011) | 0.9887(0.0013) | 0.9804(0.0013) | 0.9771(0.0015) | 0.9785(0.0020) | 0.9841(0.0018) | 0.9868(0.0007) | 0.9870(0.0008) | 0.9926(0.0007) | 0.9769(0.0008) | 0.9863(0.0022) |
| lag 09 | 0.9993(0.0019) | 0.9941(0.0011) | 0.9888(0.0013) | 0.9797(0.0013) | 0.9748(0.0016) | 0.9773(0.0022) | 0.9830(0.0019) | 0.9862(0.0007) | 0.9865(0.0009) | 0.9925(0.0007) | 0.9757(0.0009) | 0.9848(0.0024) |
| lag 010 | 0.9998(0.0020) | 0.9942(0.0012) | 0.9890(0.0014) | 0.9790(0.0014) | 0.9733(0.0016) | 0.9768(0.0023) | 0.9817(0.0020) | 0.9859(0.0008) | 0.9860(0.0010) | 0.9925(0.0008) | 0.9748(0.0010) | 0.9835(0.0025) |

**Table S6 . OR (SD) of acute surface pressure exposure and HDFM at different single lag days and moving average lag days in Jiangsu, China, 2009-2023.**

|  | Age | | | | | | | Gender | | Diagnostic type | | |
| --- | --- | --- | --- | --- | --- | --- | --- | --- | --- | --- | --- | --- |
|  | ＜1 | 1-＜2 | 2-＜3 | 3-＜4 | 4-＜5 | 5-＜6 | ≥6 | Male | Female | Laboratory confirmed | Clinically confirmed | Confirmed |
| lag 1 | 0.9889(0.0079) | 1.0057(0.0048) | 1.0211(0.0056) | 1.0458(0.0055) | 1.0537(0.0065) | 1.0440(0.0093) | 1.0184(0.0084) | 1.0179(0.0031) | 1.0344(0.0038) | 1.0044(0.0031) | 1.0589(0.0040) | 1.0170(0.0103) |
| lag 2 | 0.9911(0.0078) | 1.0112(0.0048) | 1.0334(0.0056) | 1.0641(0.0055) | 1.0774(0.0065) | 1.0615(0.0092) | 1.0379(0.0084) | 1.0313(0.0031) | 1.0470(0.0038) | 1.0134(0.0031) | 1.0780(0.0040) | 1.0377(0.0103) |
| lag 3 | 0.9757(0.0078) | 0.9884(0.0047) | 1.0204(0.0056) | 1.0535(0.0055) | 1.0802(0.0064) | 1.0504(0.0092) | 1.0303(0.0084) | 1.0177(0.0031) | 1.0353(0.0038) | 0.9961(0.0031) | 1.0717(0.0040) | 1.0364(0.0102) |
| lag 4 | 0.9326(0.0078) | 0.9498(0.0047) | 0.9721(0.0056) | 1.0185(0.0055) | 1.0396(0.0064) | 1.0049(0.0091) | 0.9968(0.0083) | 0.9769(0.0031) | 0.9956(0.0037) | 0.9547(0.0031) | 1.0306(0.0039) | 1.0106(0.0102) |
| lag 5 | 0.9076(0.0077) | 0.9130(0.0047) | 0.9324(0.0055) | 0.9803(0.0054) | 0.9894(0.0063) | 0.9699(0.0091) | 0.9502(0.0083) | 0.9383(0.0031) | 0.9565(0.0037) | 0.9210(0.0031) | 0.9833(0.0039) | 0.9676(0.0101) |
| lag 6 | 0.9029(0.0077) | 0.8934(0.0047) | 0.9112(0.0055) | 0.9472(0.0054) | 0.9530(0.0063) | 0.9545(0.0090) | 0.9223(0.0083) | 0.9187(0.0031) | 0.9272(0.0037) | 0.9024(0.0031) | 0.9519(0.0039) | 0.9396(0.0101) |
| lag 7 | 0.9114(0.0076) | 0.8945(0.0046) | 0.9096(0.0055) | 0.9407(0.0054) | 0.9448(0.0063) | 0.9453(0.0090) | 0.9216(0.0082) | 0.9204(0.0030) | 0.9191(0.0037) | 0.9008(0.0030) | 0.9491(0.0039) | 0.9354(0.0100) |
| lag 8 | 0.9146(0.0076) | 0.9018(0.0046) | 0.9144(0.0055) | 0.9529(0.0054) | 0.9585(0.0062) | 0.9570(0.0089) | 0.9381(0.0082) | 0.9300(0.0030) | 0.9276(0.0037) | 0.9048(0.0030) | 0.9672(0.0039) | 0.9456(0.0100) |
| lag 9 | 0.9213(0.0076) | 0.9108(0.0046) | 0.9222(0.0054) | 0.9642(0.0053) | 0.9645(0.0062) | 0.9613(0.0089) | 0.9455(0.0081) | 0.9366(0.0030) | 0.9381(0.0036) | 0.9116(0.0030) | 0.9782(0.0038) | 0.9490(0.0100) |
| lag 10 | 0.9338(0.0075) | 0.9327(0.0046) | 0.9366(0.0054) | 0.9705(0.0053) | 0.9747(0.0062) | 0.9655(0.0089) | 0.9556(0.0081) | 0.9506(0.0030) | 0.9498(0.0036) | 0.9286(0.0030) | 0.9844(0.0038) | 0.9627(0.0100) |
| lag 01 | 0.9823(0.0085) | 0.9972(0.0051) | 1.0086(0.0060) | 1.0350(0.0059) | 1.0482(0.0070) | 1.0328(0.0099) | 0.9985(0.0090) | 1.0069(0.0034) | 1.0255(0.0041) | 0.9940(0.0034) | 1.0497(0.0043) | 1.0025(0.0109) |
| lag 02 | 0.9826(0.0090) | 1.0027(0.0055) | 1.0213(0.0065) | 1.0553(0.0064) | 1.0716(0.0075) | 1.0524(0.0107) | 1.0153(0.0096) | 1.0190(0.0036) | 1.0402(0.0044) | 1.0013(0.0036) | 1.0730(0.0046) | 1.0181(0.0117) |
| lag 03 | 0.9762(0.0096) | 0.9979(0.0058) | 1.0258(0.0069) | 1.0676(0.0068) | 1.0923(0.0080) | 1.0642(0.0114) | 1.0242(0.0103) | 1.0229(0.0038) | 1.0477(0.0046) | 0.9996(0.0038) | 1.0901(0.0049) | 1.0289(0.0125) |
| lag 04 | 0.9557(0.0101) | 0.9808(0.0062) | 1.0132(0.0073) | 1.0669(0.0072) | 1.0969(0.0084) | 1.0590(0.0120) | 1.0204(0.0108) | 1.0123(0.0040) | 1.0410(0.0049) | 0.9840(0.0040) | 1.0916(0.0052) | 1.0293(0.0131) |
| lag 05 | 0.9300(0.0107) | 0.9541(0.0065) | 0.9899(0.0077) | 1.0549(0.0075) | 1.0853(0.0089) | 1.0438(0.0126) | 1.0025(0.0114) | 0.9909(0.0043) | 1.0232(0.0052) | 0.9597(0.0043) | 1.0783(0.0055) | 1.0164(0.0138) |
| lag 06 | 0.9050(0.0112) | 0.9241(0.0068) | 0.9629(0.0081) | 1.0344(0.0079) | 1.0645(0.0093) | 1.0265(0.0133) | 0.9784(0.0119) | 0.9661(0.0045) | 0.9986(0.0054) | 0.9322(0.0045) | 1.0577(0.0057) | 0.9971(0.0145) |
| lag 07 | 0.8837(0.0118) | 0.8962(0.0072) | 0.9372(0.0085) | 1.0144(0.0083) | 1.0439(0.0098) | 1.0082(0.0139) | 0.9561(0.0125) | 0.9435(0.0047) | 0.9737(0.0057) | 0.9058(0.0047) | 1.0390(0.0060) | 0.9782(0.0152) |
| lag 08 | 0.8642(0.0123) | 0.8719(0.0075) | 0.9146(0.0089) | 0.9997(0.0087) | 1.0296(0.0102) | 0.9949(0.0146) | 0.9400(0.0131) | 0.9251(0.0049) | 0.9531(0.0060) | 0.8820(0.0049) | 1.0277(0.0063) | 0.9636(0.0159) |
| lag 09 | 0.8479(0.0128) | 0.8517(0.0078) | 0.8955(0.0092) | 0.9891(0.0091) | 1.0180(0.0107) | 0.9834(0.0152) | 0.9266(0.0136) | 0.9096(0.0051) | 0.9367(0.0062) | 0.8618(0.0051) | 1.0203(0.0066) | 0.9505(0.0166) |
| lag 010 | 0.8362(0.0132) | 0.8387(0.0081) | 0.8814(0.0095) | 0.9808(0.0094) | 1.0099(0.0111) | 0.9738(0.0158) | 0.9166(0.0142) | 0.8988(0.0053) | 0.9244(0.0064) | 0.8474(0.0053) | 1.0151(0.0068) | 0.9415(0.0172) |

**Table S7 . OR (SD) of acute precipitation exposure and HDFM at different single lag days and moving average lag days in Jiangsu, China, 2009-2023.**

|  | Age | | | | | | | Gender | | Diagnostic type | | |
| --- | --- | --- | --- | --- | --- | --- | --- | --- | --- | --- | --- | --- |
|  | ＜1 | 1-＜2 | 2-＜3 | 3-＜4 | 4-＜5 | 5-＜6 | ≥6 | Male | Female | Laboratory confirmed | Clinically confirmed | Confirmed |
| lag 1 | 1.0002(0.0006) | 0.9997(0.0004) | 0.9994(0.0005) | 0.9988(0.0005) | 1.0005(0.0006) | 0.9995(0.0008) | 1.0018(0.0007) | 0.9999(0.0003) | 0.9997(0.0003) | 0.9997(0.0003) | 0.9996(0.0004) | 1.0026(0.0008) |
| lag 2 | 1.0006(0.0006) | 1.0004(0.0004) | 1.0005(0.0005) | 0.9993(0.0005) | 1.0006(0.0006) | 0.9993(0.0008) | 1.0014(0.0006) | 1.0003(0.0003) | 1.0003(0.0003) | 1.0003(0.0003) | 1.0000(0.0004) | 1.0012(0.0008) |
| lag 3 | 1.0014(0.0007) | 1.0004(0.0004) | 1.0003(0.0005) | 0.9999(0.0005) | 0.9995(0.0006) | 1.0014(0.0008) | 1.0002(0.0007) | 1.0002(0.0003) | 1.0005(0.0003) | 1.0003(0.0003) | 1.0003(0.0004) | 1.0001(0.0009) |
| lag 4 | 1.0012(0.0006) | 1.0004(0.0004) | 1.0014(0.0005) | 1.0023(0.0005) | 1.0029(0.0006) | 1.0036(0.0008) | 1.0014(0.0007) | 1.0016(0.0003) | 1.0015(0.0003) | 1.0012(0.0003) | 1.0026(0.0004) | 1.0006(0.0009) |
| lag 5 | 1.0013(0.0006) | 1.0012(0.0004) | 1.0020(0.0005) | 1.0023(0.0005) | 1.0036(0.0006) | 1.0049(0.0008) | 1.0032(0.0007) | 1.0024(0.0003) | 1.0021(0.0003) | 1.0013(0.0003) | 1.0042(0.0004) | 1.0026(0.0009) |
| lag 6 | 1.0018(0.0006) | 1.0016(0.0004) | 1.0020(0.0005) | 1.0041(0.0005) | 1.0047(0.0006) | 1.0040(0.0008) | 1.0036(0.0007) | 1.0030(0.0003) | 1.0026(0.0003) | 1.0018(0.0003) | 1.0051(0.0004) | 1.0023(0.0009) |
| lag 7 | 1.0017(0.0006) | 1.0020(0.0004) | 1.0029(0.0005) | 1.0040(0.0005) | 1.0050(0.0006) | 1.0031(0.0008) | 1.0027(0.0007) | 1.0030(0.0003) | 1.0029(0.0003) | 1.0024(0.0003) | 1.0046(0.0004) | 1.0008(0.0009) |
| lag 8 | 1.0001(0.0007) | 1.0008(0.0004) | 1.0015(0.0005) | 1.0009(0.0005) | 1.0014(0.0006) | 1.0007(0.0008) | 1.0008(0.0007) | 1.0009(0.0003) | 1.0011(0.0003) | 1.0010(0.0003) | 1.0009(0.0004) | 1.0008(0.0009) |
| lag 9 | 1.0000(0.0007) | 1.0006(0.0004) | 1.0007(0.0005) | 0.9995(0.0005) | 0.9997(0.0006) | 1.0008(0.0008) | 1.0004(0.0007) | 1.0004(0.0003) | 1.0001(0.0003) | 1.0006(0.0003) | 0.9994(0.0004) | 1.0012(0.0009) |
| lag 10 | 1.0002(0.0007) | 1.0003(0.0004) | 1.0011(0.0005) | 1.0012(0.0005) | 0.9988(0.0006) | 1.0003(0.0008) | 1.0015(0.0007) | 1.0004(0.0003) | 1.0007(0.0003) | 1.0006(0.0003) | 1.0000(0.0004) | 1.0019(0.0009) |
| lag 01 | 0.9984(0.0008) | 0.9988(0.0005) | 0.9982(0.0006) | 0.9971(0.0006) | 0.9987(0.0007) | 0.9976(0.0010) | 1.0019(0.0008) | 0.9985(0.0003) | 0.9986(0.0004) | 0.9985(0.0003) | 0.9978(0.0005) | 1.0030(0.0011) |
| lag 02 | 0.9990(0.0010) | 0.9991(0.0006) | 0.9987(0.0007) | 0.9968(0.0007) | 0.9992(0.0009) | 0.9972(0.0012) | 1.0028(0.0010) | 0.9989(0.0004) | 0.9989(0.0005) | 0.9989(0.0004) | 0.9980(0.0005) | 1.0037(0.0013) |
| lag 03 | 1.0000(0.0011) | 0.9994(0.0006) | 0.9990(0.0008) | 0.9968(0.0008) | 0.9989(0.0010) | 0.9983(0.0013) | 1.0029(0.0011) | 0.9991(0.0004) | 0.9993(0.0005) | 0.9992(0.0004) | 0.9982(0.0006) | 1.0036(0.0014) |
| lag 04 | 1.0009(0.0012) | 0.9997(0.0007) | 1.0000(0.0009) | 0.9985(0.0009) | 1.0009(0.0011) | 1.0008(0.0015) | 1.0039(0.0012) | 1.0002(0.0005) | 1.0003(0.0006) | 1.0000(0.0005) | 1.0000(0.0007) | 1.0039(0.0016) |
| lag 05 | 1.0018(0.0013) | 1.0005(0.0008) | 1.0013(0.0010) | 1.0000(0.0010) | 1.0034(0.0012) | 1.0041(0.0016) | 1.0059(0.0014) | 1.0018(0.0005) | 1.0017(0.0006) | 1.0009(0.0005) | 1.0028(0.0008) | 1.0056(0.0017) |
| lag 06 | 1.0030(0.0014) | 1.0015(0.0008) | 1.0026(0.0010) | 1.0027(0.0011) | 1.0067(0.0013) | 1.0069(0.0017) | 1.0085(0.0015) | 1.0038(0.0006) | 1.0034(0.0007) | 1.0021(0.0006) | 1.0064(0.0008) | 1.0072(0.0019) |
| lag 07 | 1.0042(0.0015) | 1.0030(0.0009) | 1.0048(0.0011) | 1.0057(0.0012) | 1.0107(0.0014) | 1.0095(0.0019) | 1.0107(0.0016) | 1.0061(0.0006) | 1.0056(0.0008) | 1.0039(0.0006) | 1.0100(0.0009) | 1.0081(0.0021) |
| lag 08 | 1.0045(0.0017) | 1.0037(0.0010) | 1.0061(0.0012) | 1.0066(0.0013) | 1.0123(0.0015) | 1.0104(0.0021) | 1.0118(0.0017) | 1.0070(0.0007) | 1.0067(0.0008) | 1.0047(0.0007) | 1.0112(0.0010) | 1.0091(0.0022) |
| lag 09 | 1.0047(0.0018) | 1.0042(0.0011) | 1.0068(0.0013) | 1.0065(0.0014) | 1.0126(0.0016) | 1.0116(0.0022) | 1.0127(0.0019) | 1.0076(0.0007) | 1.0070(0.0009) | 1.0054(0.0007) | 1.0112(0.0010) | 1.0104(0.0024) |
| lag 010 | 1.0050(0.0019) | 1.0046(0.0011) | 1.0079(0.0014) | 1.0077(0.0015) | 1.0122(0.0018) | 1.0123(0.0024) | 1.0145(0.0020) | 1.0082(0.0008) | 1.0078(0.0009) | 1.0061(0.0008) | 1.0116(0.0011) | 1.0124(0.0026) |

**Table S8 . Two-meteorological models showing associations of 10-d moving average (lag010) exposures to temperature, humidity, wind speed, radiation, surface pressure, and precipitation with HDFM after adjustment for other meteorological factors in Jiangsu, China, 2009-2023.**

| Primary exposure | Adjusted | OR (95% CI) |
| --- | --- | --- |
| Temperature | Humidity | 1.0104 (1.0093, 1.0115) |
|  | Wind speed | 1.0176 (1.0165, 1.0187) |
|  | Radiation | 1.0123 (1.0113, 1.0134) |
|  | Surface pressure | 1.0069 (1.0054, 1.0083) |
|  | Precipitation | 1.0129 (1.0118, 1.0139) |
| Humidity | Temperature | 1.0057 (1.0053, 1.0061) |
|  | Wind speed | 1.0051 (1.0045, 1.0057) |
|  | Radiation | 1.0064 (1.006, 1.0069) |
|  | Surface pressure | 1.0052 (1.0048, 1.0057) |
|  | Precipitation | 1.0063 (1.0058, 1.0068) |
| Wind speed | Temperature | 1.0047 (1.0001, 1.0094) |
|  | Humidity | 1.0134 (1.0087, 1.0182) |
|  | Radiation | 1.0082 (1.0035, 1.0129) |
|  | Surface pressure | 1.0052 (1.0005, 1.0099) |
|  | Precipitation | 1.0076 (1.0029, 1.0123) |
| Radiation | Temperature | 0.9803 (0.9791, 0.9815) |
|  | Humidity | 0.9949 (0.9934, 0.9965) |
|  | Wind speed | 0.9859 (0.9847, 0.9871) |
|  | Surface pressure | 0.9856 (0.9844, 0.9868) |
|  | Precipitation | 0.9858 (0.9844, 0.9872) |
| Surface pressure | Temperature | 0.9428 (0.9323, 0.9534) |
|  | Humidity | 0.9342 (0.9265, 0.942) |
|  | Wind speed | 0.9067 (0.8995, 0.9139) |
|  | Radiation | 0.9092 (0.902, 0.9165) |
|  | Precipitation | 0.9151 (0.9077, 0.9225) |
| Precipitation | Temperature | 1.009 (1.0078, 1.0102) |
|  | Humidity | 1.0004 (0.9991, 1.0017) |
|  | Wind speed | 0.9997 (0.9983, 1.0012) |
|  | Radiation | 1.0081 (1.0069, 1.0093) |
|  | Surface pressure | 1.0058 (1.0046, 1.007) |

**Table S9. OR (SD) of acute meteorological exposure and HDFM at different single lag days and moving average lag days when restricting analyses in Jiangsu, China, 2009-2019.**

|  | Temperature | Humidity | Wind speed | Radiation | Surface pressure | Precipitation |
| --- | --- | --- | --- | --- | --- | --- |
| Lag 1 | 0.9957 (0.0004) | 0.9988 (0.0001) | 0.9922 (0.0010) | 1.0017 (0.0003) | 1.0249 (0.0026) | 0.9995 (0.0002) |
| Lag 2 | 0.9921 (0.0004) | 0.9987 (0.0001) | 0.9976 (0.0010) | 1.0002 (0.0003) | 1.0376 (0.0026) | 0.9999 (0.0002) |
| Lag 3 | 0.9929 (0.0004) | 0.9997 (0.0001) | 1.0048 (0.0010) | 0.9975 (0.0003) | 1.0239 (0.0026) | 1.0002 (0.0002) |
| Lag 4 | 0.9993 (0.0004) | 1.0015 (0.0001) | 1.0053 (0.0010) | 0.9948 (0.0003) | 0.9815 (0.0026) | 1.0016 (0.0002) |
| Lag 5 | 1.0058 (0.0004) | 1.0032 (0.0001) | 1.0051 (0.0010) | 0.9928 (0.0003) | 0.9417 (0.0026) | 1.0024 (0.0002) |
| Lag 6 | 1.0101 (0.0004) | 1.0041 (0.0001) | 1.0063 (0.0010) | 0.9925 (0.0003) | 0.9170 (0.0026) | 1.0030 (0.0002) |
| Lag 7 | 1.0119 (0.0004) | 1.0036 (0.0001) | 1.0089 (0.0010) | 0.9947 (0.0003) | 0.9126 (0.0025) | 1.0034 (0.0002) |
| Lag 8 | 1.0121 (0.0004) | 1.0022 (0.0001) | 1.0075 (0.0010) | 0.9976 (0.0003) | 0.9199 (0.0025) | 1.0014 (0.0002) |
| Lag 9 | 1.0113 (0.0004) | 1.0014 (0.0001) | 1.0066 (0.0010) | 0.9996 (0.0003) | 0.9293 (0.0025) | 1.0007 (0.0002) |
| Lag 10 | 1.0086 (0.0004) | 1.0010 (0.0001) | 1.0030 (0.0010) | 1.0005 (0.0003) | 0.9455 (0.0025) | 1.0005 (0.0002) |
| Lag 01 | 0.9994 (0.0004) | 0.9984 (0.0001) | 0.9852 (0.0013) | 1.0048 (0.0003) | 1.0164 (0.0028) | 0.9979 (0.0003) |
| Lag 02 | 0.9962 (0.0004) | 0.9980 (0.0001) | 0.9854 (0.0014) | 1.0043 (0.0004) | 1.0292 (0.0030) | 0.9980 (0.0003) |
| Lag 03 | 0.9943 (0.0005) | 0.9981 (0.0002) | 0.9894 (0.0016) | 1.0026 (0.0004) | 1.0340 (0.0032) | 0.9982 (0.0004) |
| Lag 04 | 0.9947 (0.0005) | 0.9989 (0.0002) | 0.9930 (0.0017) | 0.9996 (0.0004) | 1.0237 (0.0034) | 0.9993 (0.0004) |
| Lag 05 | 0.9969 (0.0005) | 1.0003 (0.0002) | 0.9960 (0.0019) | 0.9960 (0.0005) | 1.0022 (0.0036) | 1.0009 (0.0005) |
| Lag 06 | 0.9999 (0.0005) | 1.0020 (0.0002) | 0.9996 (0.0020) | 0.9924 (0.0005) | 0.9754 (0.0038) | 1.0029 (0.0005) |
| Lag 07 | 1.0030 (0.0005) | 1.0035 (0.0002) | 1.0047 (0.0022) | 0.9897 (0.0005) | 0.9494 (0.0040) | 1.0053 (0.0005) |
| Lag 08 | 1.0060 (0.0006) | 1.0045 (0.0002) | 1.0094 (0.0024) | 0.9884 (0.0006) | 0.9275 (0.0042) | 1.0066 (0.0006) |
| Lag 09 | 1.0085 (0.0006) | 1.0052 (0.0002) | 1.0139 (0.0025) | 0.9880 (0.0006) | 0.9097 (0.0043) | 1.0073 (0.0006) |
| Lag 010 | 1.0102 (0.0006) | 1.0057 (0.0002) | 1.0162 (0.0027) | 0.9880 (0.0007) | 0.8976 (0.0045) | 1.0080 (0.0007) |

**Table S10. OR (SD) of acute meteorological exposure and HDFM at different single lag days and moving average lag days when restricting analyses in Jiangsu, China, 2020-2023.**

|  | Temperature | Humidity | Wind speed | Radiation | Surface pressure | Precipitation |
| --- | --- | --- | --- | --- | --- | --- |
| Lag 1 | 1.0026 (0.0008) | 1.0005 (0.0003) | 0.9930 (0.0021) | 0.9959 (0.0006) | 1.0194 (0.0057) | 1.0010 (0.0004) |
| Lag 2 | 0.9999 (0.0008) | 0.9994 (0.0003) | 0.9958 (0.0021) | 0.9962 (0.0006) | 1.0393 (0.0057) | 1.0015 (0.0004) |
| Lag 3 | 1.0029 (0.0008) | 0.9998 (0.0003) | 1.0068 (0.0021) | 0.9952 (0.0006) | 1.0334 (0.0057) | 1.0003 (0.0004) |
| Lag 4 | 1.0088 (0.0008) | 1.0019 (0.0003) | 1.0149 (0.0022) | 0.9924 (0.0006) | 1.0003 (0.0057) | 1.0012 (0.0004) |
| Lag 5 | 1.0136 (0.0008) | 1.0035 (0.0003) | 1.0062 (0.0022) | 0.9930 (0.0006) | 0.9633 (0.0057) | 1.0020 (0.0005) |
| Lag 6 | 1.0153 (0.0008) | 1.0042 (0.0003) | 0.9985 (0.0022) | 0.9932 (0.0006) | 0.9451 (0.0057) | 1.0028 (0.0005) |
| Lag 7 | 1.0148 (0.0008) | 1.0039 (0.0003) | 0.9981 (0.0023) | 0.9942 (0.0006) | 0.9568 (0.0057) | 1.0017 (0.0004) |
| Lag 8 | 1.0157 (0.0008) | 1.0022 (0.0003) | 0.9990 (0.0022) | 0.9984 (0.0006) | 0.9769 (0.0057) | 0.9994 (0.0004) |
| Lag 9 | 1.0150 (0.0008) | 1.0021 (0.0003) | 0.9946 (0.0022) | 0.9985 (0.0006) | 0.9795 (0.0057) | 0.9987 (0.0004) |
| Lag 10 | 1.0119 (0.0008) | 1.0029 (0.0003) | 0.9910 (0.0022) | 0.9960 (0.0006) | 0.9776 (0.0057) | 1.0010 (0.0004) |
| Lag 01 | 1.0065 (0.0009) | 1.0009 (0.0003) | 0.9888 (0.0025) | 0.9951 (0.0007) | 0.9998 (0.0061) | 1.0015 (0.0006) |
| Lag 02 | 1.0049 (0.0010) | 1.0004 (0.0003) | 0.9881 (0.0028) | 0.9936 (0.0008) | 1.0166 (0.0065) | 1.0026 (0.0007) |
| Lag 03 | 1.0053 (0.0010) | 1.0003 (0.0004) | 0.9925 (0.0031) | 0.9916 (0.0009) | 1.0264 (0.0069) | 1.0027 (0.0008) |
| Lag 04 | 1.0075 (0.0011) | 1.0012 (0.0004) | 0.9997 (0.0033) | 0.9881 (0.0010) | 1.0236 (0.0073) | 1.0036 (0.0009) |
| Lag 05 | 1.0108 (0.0011) | 1.0027 (0.0004) | 1.0025 (0.0036) | 0.9848 (0.0011) | 1.0103 (0.0077) | 1.0050 (0.0009) |
| Lag 06 | 1.0144 (0.0012) | 1.0045 (0.0005) | 1.0018 (0.0038) | 0.9814 (0.0012) | 0.9938 (0.0081) | 1.0071 (0.0010) |
| Lag 07 | 1.0175 (0.0012) | 1.0063 (0.0005) | 1.0010 (0.0041) | 0.9782 (0.0013) | 0.9822 (0.0085) | 1.0088 (0.0011) |
| Lag 08 | 1.0208 (0.0012) | 1.0073 (0.0005) | 1.0006 (0.0044) | 0.9772 (0.0014) | 0.9762 (0.0089) | 1.0089 (0.0012) |
| Lag 09 | 1.0236 (0.0013) | 1.0084 (0.0006) | 0.9982 (0.0047) | 0.9761 (0.0014) | 0.9709 (0.0094) | 1.0084 (0.0013) |
| Lag 010 | 1.0255 (0.0013) | 1.0098 (0.0006) | 0.9940 (0.0050) | 0.9736 (0.0015) | 0.9653 (0.0098) | 1.0100 (0.0015) |
